# Time scale separation in the vector borne disease model SIRUV via center manifold analysis

**DOI:** 10.1101/2021.04.06.21254992

**Authors:** Maíra Aguiar, Bob Kooi, Andrea Pugliese, Mattia Sensi, Nico Stollenwerk

## Abstract

We investigate time scale separation in the vector borne disease model SIRUV, as previously described in the literature [1], and recently reanalyzed with the singular perturbation technique [2]. We focus on the analysis with a single small parameter, the birth and death rate *µ*, whereas all other model parameters are much larger and describe fast transitions. The scaling of the endemic stationary state, the Jacobian matrix around it and its eigenvalues with this small parameter *µ* is calculated and the center manifold analysis performed with the method described in [3] which goes back to earlier work [4, 5], namely a transformation of the Jacobian matrix to block structure in zeroth order in the parameter *µ* is used and then a family of center manifolds with *µ* larger than zero is obtained.

## 1 Introduction

Here we investigate the time scale separation of the fast infected mosquito dynamics *V* from the slow human infection *I* in the SIRUV model in respect to its only small parameter *µ*, describing the slow transition of building up susceptibles *S* from the recovered *R*, either by waning immunity or here via death of any human and birth of susceptibles.

The SIRUV model has been described in detail in [1] and recently investigated further e.g. in [2]. However, the full SIRUV model does not have the standard form for time scale separation, where the standard form is a separation into slow variable dynamics *dx/dt* = *εf* (*x, y, ε*) and fast dynamics *dy/dt* = *g*(*x, y, ε*) with a small scaling parameter *ε*, eventually originating from scaling of several biological model parameters. In [1] from the SIRUV a simplified model, the SISUV model, was constructed by using nearly all parameters as in the SIRUV model, but without a recovered class *R*, and therefore an extended period of infection, hence slow recovery, i.e. small *γ*, and consequently also small infectivity *β*. Hence the small parameter *ε* originates from 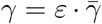 and 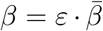 with 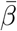 etc. in the range of the fast mosquito parameters. This simplified SISUV model has the standard form for time scale separation, and hence is a good test bed for standard techniques in comparison, such as center manifold analysis in comparison with classical time scale separation arguments like scaling of the time parameter with *ε* to obtain quasistationary expressions *V* = *V* (*I*), all this in zeroth order implicitly [1] or more recently a rigorous analysis with singular perturbation techniques beyond the zeroth order approximation [2].

But in the full SIRUV model we only have one biologically small parameter *µ* inside the slow dynamics function *f* = *f* (*x, y, µ*) and hence slow dynamics *dx/dt* = *f* (*x, y, µ*) and fast dynamics *dy/dt* = *g*(*x, y*). For such harder problems of time scale separation in the literature other techniques like Implicit Low Dimensional Manifolds (ILDM) or zero-derivative principle for slow–fast dynamical systems have been suggested and compared with each other [6]. The ILDM e.g. relies on the Jacobian matrix around any point in state space and the condition for slow manifold is given by the expression of the dynamics in eigenvector basis.

In standard time scale separation systems the comparison of ILDM and singular perturbation have shown that they agree only in lower order approximation up to second order (𝒪 (*ε*^2^)), but then deviate [7]. And such methodes like ILDM or zero-derivative principle are in danger of picking up spurious manifolds not agreeing with the Fenichel manifold of singular perturbation, in cases when they can be compared [6].

Here the center maifold analysis for the SIRUV model is presented as closer to the singular perturbation theory (and actually in a careful analysis of the scaling with small parameter the center manifold and singular perturbation agree in standard time scale separation systems, see the SISUV model again as example). Actually, a singular perturbation analysis can be performed in the SIRUV model by introducing next to the naturally small parameter *µ* a second parameter *ε* in the way of *dx/dt* = *εf* (*x, y, µ*) and *dy/dt* = *g*(*x, y*) where *ε* is not directly originating from the model parameters, but based on dimensionality arguments originating from the eigenvalue structure of the Jacobian matrix around the endemic fixed point [2]. Then *ε* has to be considered as small in respect to the mortality of the mosquitoes *ν*, compared to the mortality of the humans *µ*, but is not given directly from the model parameters and the dimensionality question has to be dealt with via extra arguments. However, the singular perturabtion gives fast calculations of high orders of the Fenichel manifold, and is therefore attractive for quick analytical treatment.

Here, in the center manifold analysis a second small parameter is not needed, and a series expansion can be obtained for the naturally small parameter *µ* and for the slow state space variables *x* giving the fast variable as manifold 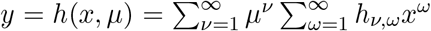, where *x* can be higher dimensional. The number of slow versus fast variables in all these methods is determined by a spectral gap, i.e. eigenvalues with small real part for the slow variables and such with much larger real part for the fast contraction directions in state space, see e.g. [6].

Another scaling has been suggested, based on the stationary states *S*\*, *I*^*\**^ and *V* *, i.e. *I* = *µĪ* and 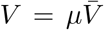 with 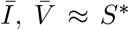 (Trento scaling), instead of the scaling from the spectral gap in the eigenvalues of the Jacobian matrix around the endemic stationary state. This scaling of state variables, however, describes a slow-fast dynamics of slow susceptibles *S* and fast infected humans *I* and infected mosquitoes *V* in the transient behaviour, before the system finally enters into the slow two-dimensional subspace of *V* = *V* (*S, I*) with *S* and *I* slow and only *V* fast, as observed from the scaling of the eigenvalues around the endemic stationary state.

## 2 The SIRUV model revisited

The SIRUV model (see [1] for a detailed description) reads as follows

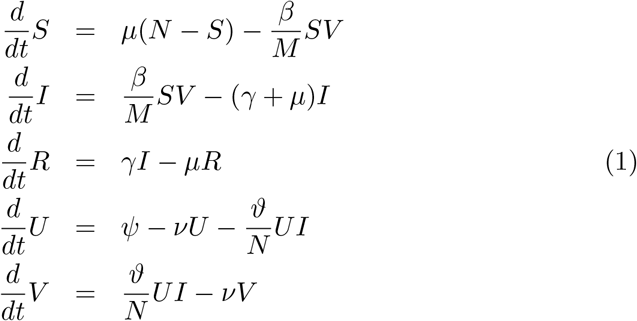

with human population size *N* = *S*(*t*)+*I*(*t*)+*R*(*t*) and mosquito population size *M* = *U* (*t*) + *V* (*t*) assumed constant, hence for now *ψ* := *ν · M*. In cosiderations of seasonality of mosquito abundance *ψ* could become seasonally forced [11].

For the human population we have birth and death rate of 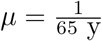, recovery rate for e.g. dengue fever of around 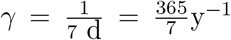, and infection rate *β* = 2 *· γ*. And for the mosquitoes life expectancy of adult mosquitoes, since only female mosquitoes bite humans for their egg production, we have 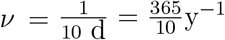, birth rate for a stable population *ψ* = *ν · M* and infection rate *ϑ* = 2 *· ν* [1].

We observe that we have only one slow parameter, the human life span or supply of new susceptibles, 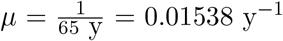. All other parameters are fast, since we have 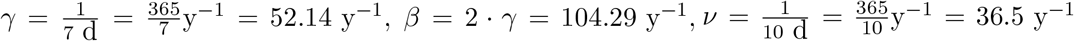 and *ϑ* = 2 *· ν* = 73.0 y^−1^. For any numerical analysis we might use as population sizes *N* = 10^6^ and *M* = 10 *· N*, hence the ratio of mosquitos to humans is *κ* = 10.

Due to the conservation of population sizes we can reduce the SIRUV model to e.g. an SIV model given by

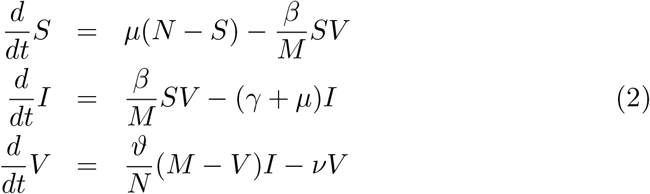

as considered recently [2]. Previously, we investigated an SRV version of the SIRUV model, since *S* and *R* both are in stationarity macroscopic variables, whereas *V* is of order *µ*, hence small and decreasing rapidly with decreasing *µ* [1]. But the biology, of course, does not change with the variables chosen for the analysis.

### 2.1 The endemic stationary state of the SIRUV model

The endemic stationary state of the SIRUV model is given by

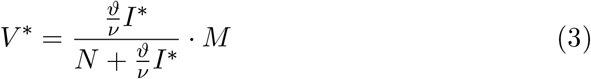

for the infected mosquitoes *V*, and

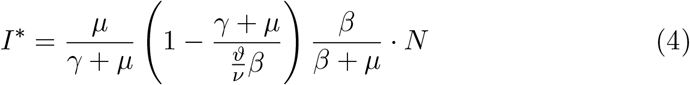

for the infected humans *I*, and

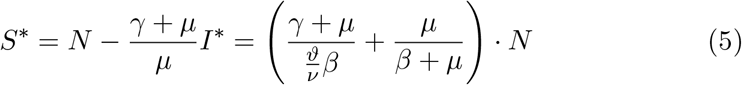

for the susceptible humans. The other variables *R* and *U* follow from the conservations, hence *R*\* = *N* − *S*\* − *I*\* and *U*\* = *M* − *V* *.

Of special interest for the following is the fact that *I* and with it also *V* scale with the small parameter *µ*, hence are of order 𝒪 (*µ*), whereas *S*\* is of order 𝒪 (1), see Eqs. (88), (89) and (90) below.

### 2.2 Stability analysis around the endemic stationary state of the SIRUV model

With the notation

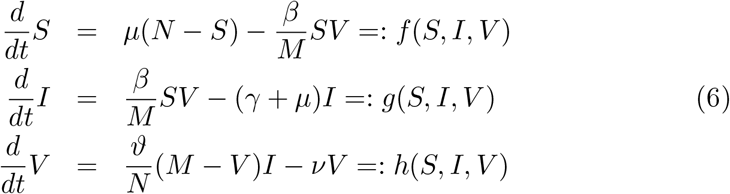

we calculate the Jacobian matrix of the SIRUV model around the endemic stationary state

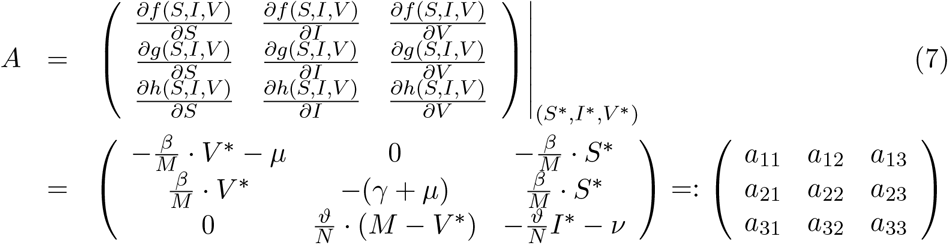

and calculate the eigenvalues of *A*. The characteristic polynomial to calculate the eigenvalues is given by

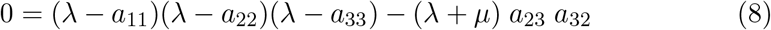

due to the structure of the Jacobian matrix *A* with e.g. *a*_13_ = −*a*_23_ and *a*_11_ = −*a*_21_ − *µ*. Multiplying out we obtain the form

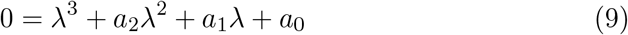

with

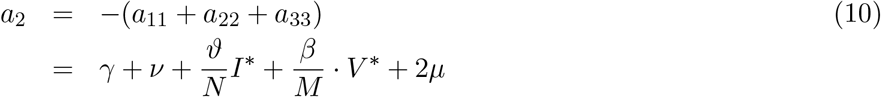

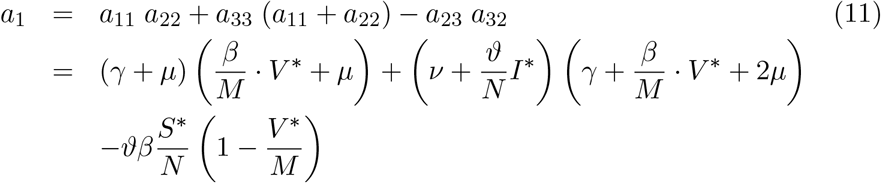

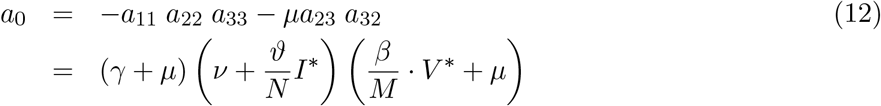

Now we can solve the characteristic polynomial of third order in *λ* with Cardano’s method, see Appendix A for the detailed analysis.

We expect a structure of the eigenvalues as follows: We have two complex eigenvalues *λ*_1*/*2_ = *a ± i · b* for the rotating part of the trajectory into the fixed point and one real eigenvalue *λ*_3_ = *c*, with *a, b* and *c* real numbers [1].

From Cardano’s method we obtain the eigenvalues numerically, see Appendix A, as

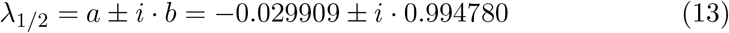

and

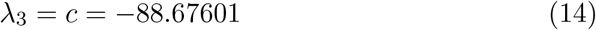

in good numerical agreement with [1], where the SRV submodel was used, and not as here the SIV submodel.

And the scaling analysis gives the largely negative real eigenvalue as

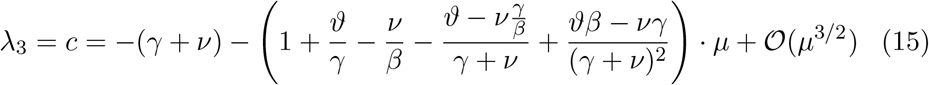

with an order 𝒪 (1) leading part and the pair of complex conjugate eigenvalues *λ*_1*/*2_ = *a±ib* with real part *a* of order 𝒪 (*µ*) and complex part *b* of order 𝒪 (*µ*^1*/*2^) explicitly as

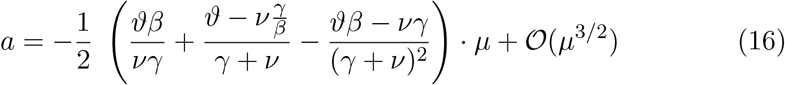

and

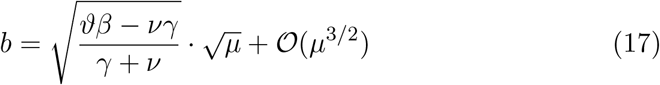

with the detailed calculations given in Appendix A.

### 2.3 Qualitative behaviour of the SIRUV model near the endemic stationary state

Since we analyse the long term dynamics into the endemic fixed point, we show here trajectories with initial conditions close to the fixed point. In Fig. 1 a) we observe in the time series of infected humans *I* oscillations into the fixed point given by the straight line. The state space plot of infected humans *I* versus susceptible humans *S* in Fig. 1 b) shows after a brief transient parallel to the y-axis, hence fast decreasing numbers of infected the spiraling into the fixed point with little non-linear deformation left.

**Figure 1:**
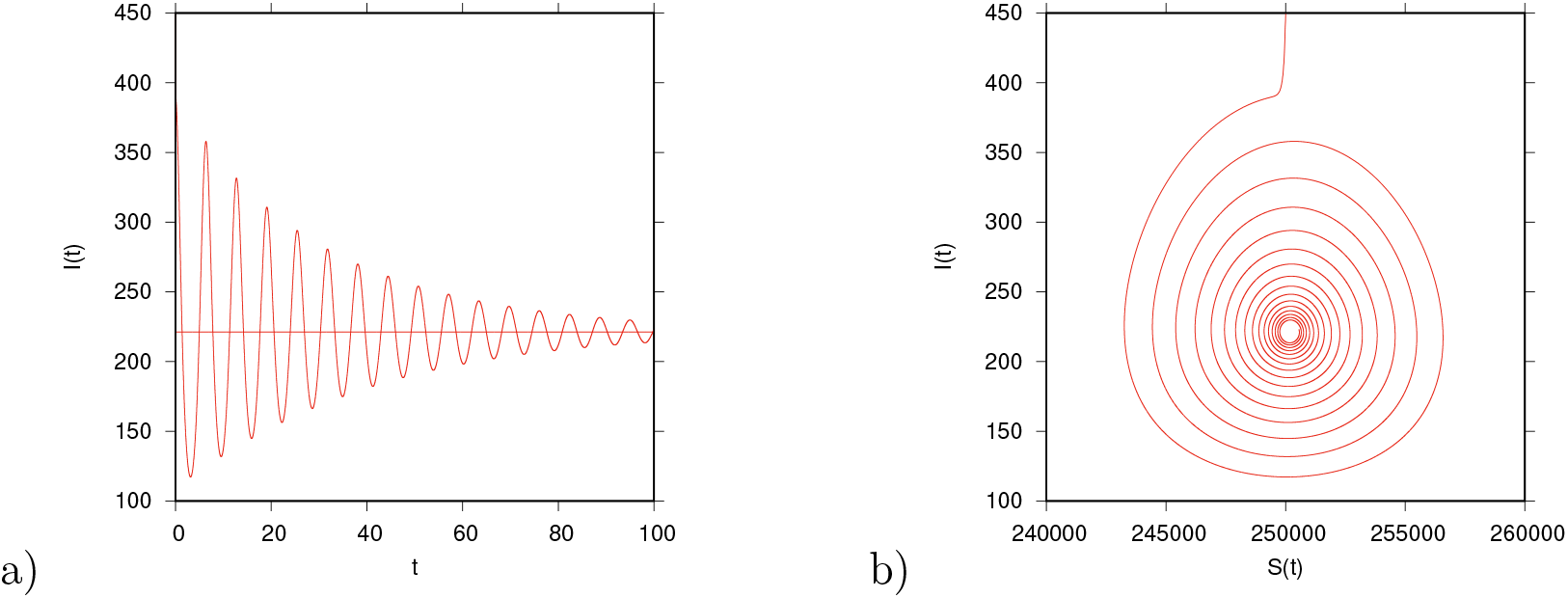
a) Time series of infected and b) state space plot of infected versus susceptibles in the SIRUV model with initial conditions close to the endemic fixed point. After an initial transient of decreasing infected I, oscillations symmetrically around the fixed point are observed.

For the infected mosquitoes *V* the time series in Fig. 2 a) shows a similar dynamics of oscillations into the fixed point as the human infected *I* in Fig. 1 a). In addition the state space plot in Fig. 2 b) reveals, after a brief transient, a close to linear functional relation between *V* and *I*, namely

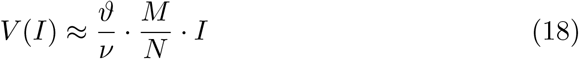

and with the parameter values used here for the SIRUV model, we obtain in good approximation *V* (*I*) ≈ 2 *·* 10 *· I*(*t*). There is only a minor tilting visible to just observe a spiraling in the complete three dimensional state space of *S, I* and *V* from this two dimensional projection of *I* and *V* only.

**Figure 2:**
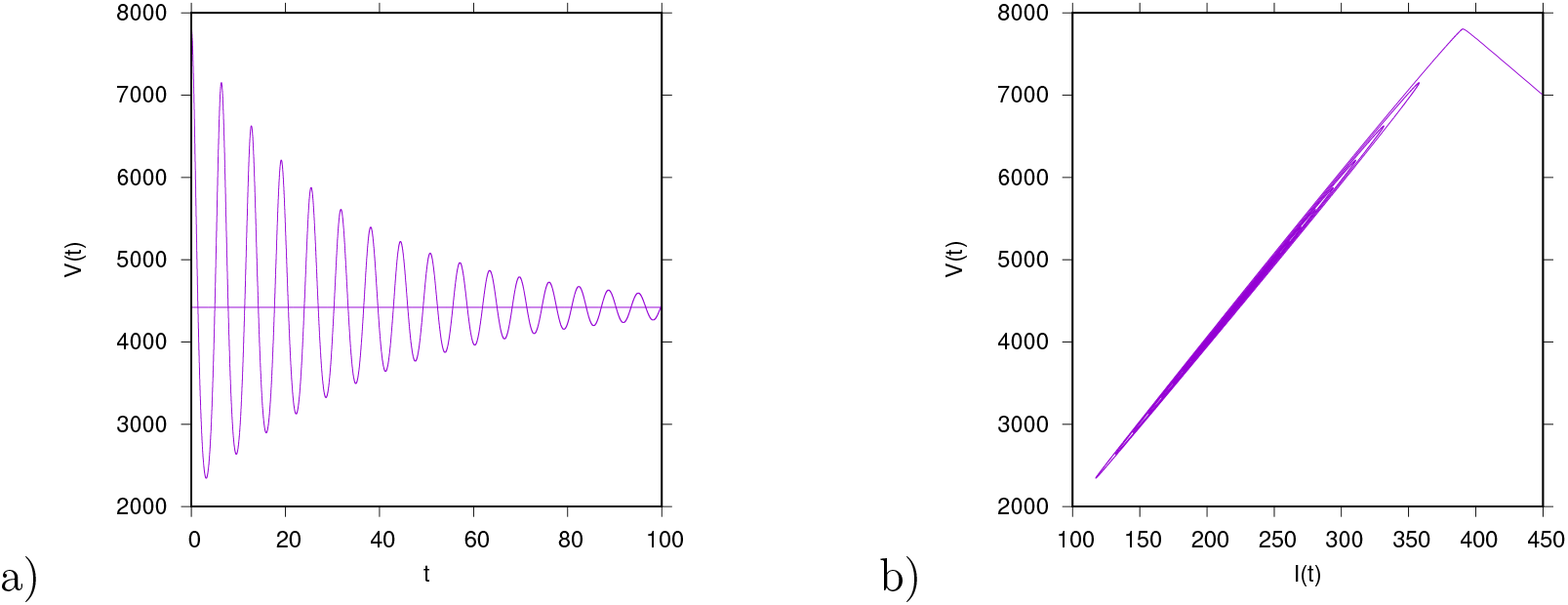
a) Time series of infected mosquitoes and b) state space plot of infected humans versus infected mosquitoes in the SIRUV model with initial conditions close to the endemic fixed point. The infected mosquitoes follow after a short transient period the infected humans with 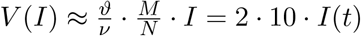

For more information in larger regions of the state space see graphs in [2], as well as here in SIV state space formulation, and similarly for the SRV state space formulation of the SIRUV model in [1].

## 3 Center manifold analysis of the SIRUV model

### 3.1 General ansatz of the center manifold analysis of the SIRUV

For the further analysis of the center manifold we first transform the dynamical system so that the endemic fixed point is in the center of the coordinate system.

The original SIRUV system is given by

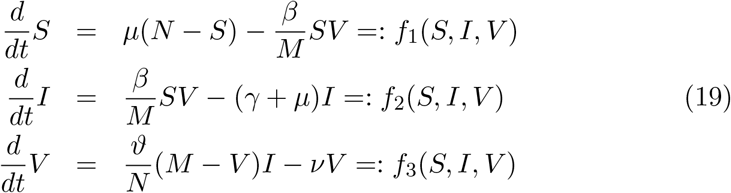

and with centered coordinates

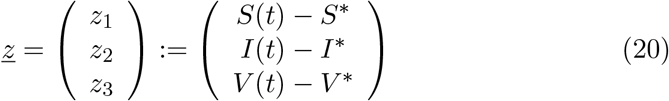

and hence for the original state variables

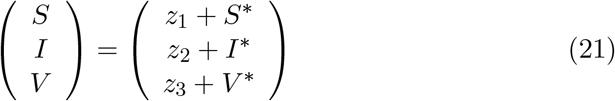

we obtain

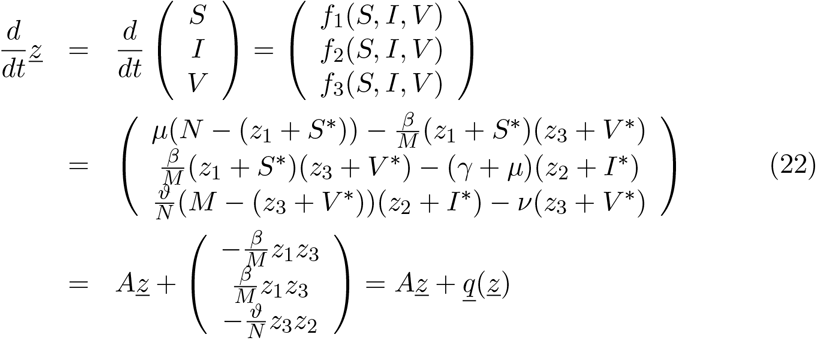

with *A* the Jacobian matrix of the SIRUV model around the endemic stationary state and *<u>q</u>*(*<u>z</u>*) the nonlinear part of the dynamics. Hence we have in centered coordinates the system given by

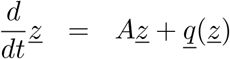

with the non-linear parts calculated as

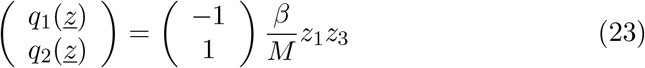

and

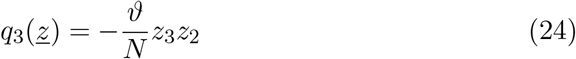

Then with the knowledge of the eigenvalues and its scaling we could calculate the, in this case, complex eigenvectors *<u>u</u>*_*i*_ and with the transformation matrix

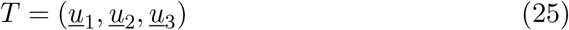

and the transformation *<u>z</u>* = *T<u>x</u>*_3_ we obtain the diagonalized system with eigenvalue matrix Λ in the form

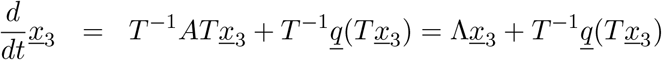

The components of the in this case 3-dimensional vector *<u>x</u>*_3_ are in time scale analysis typically grouped as

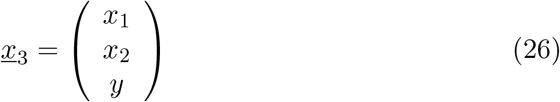

since the first two eigenvalues in Λ, *λ*_1*/*2_ = *a ± ib*, are small, corresponding to the state variables *<u>x</u>* := (*x*_1_, *x*_2_)^*tr*^ in the transformed diagonalized system as slow variables, and the third eigenvalue is largely negative, *λ*_3_ = *c*, corresponding to the state variables *y* in the transformed diagonalized system as fast variable.

Hence we then have the system separated into fast and slow variables given as

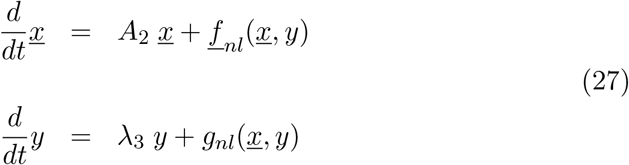

because of the spectral gap between the small *λ*_1*/*2_ = *a±ib* and the largely negative *λ*_3_ = *c*. The nonlinear parts *<u>f</u>*_*nl*_(*<u>x</u>, y*) and *g*_*nl*_(*<u>x</u>, y*) have to be determined from *T*^−1^*<u>q</u>*(*T<u>x</u>*_3_).

Here in place of *A*_2_ with index for a 2 dimensional matrix, we could simply set the matrix of the two eigenvalues

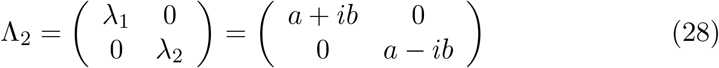

or, instead of using complex eigenvectors, we can set a real matrix, which has the same eigenvalues as Λ_2_, e.g. as

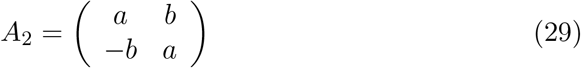

Then the full 3 dimensional matrix A_3_ in block structure is given by

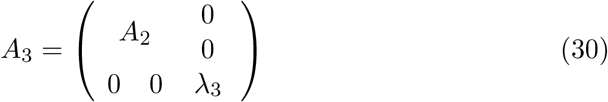

Then the center manifold is given by the fast variable *y* as a function of the slow variables *<u>x</u>* := (*x*_1_, *x*_2_)^*tr*^

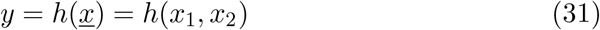

where *h* is now in the SIRUV model a vector valued function and the functional equation 𝒩 (*h*(*<u>x</u>*)) = 0 becomes

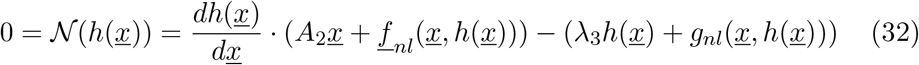

with

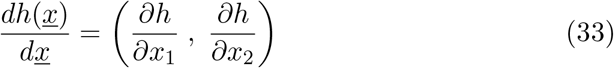

Solutions of *h*(*<u>x</u>*) can often be found by series expansion of *h* in its variables *x*_1_ and *x*_2_, and since the constant and the linear part are already dealt with by the previous transformation one can start with quadratic terms, hence the ansatz

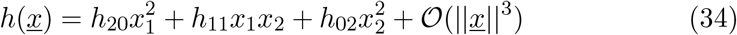

to be inserted into the equation 𝒩 (*h*(*<u>x</u>*)) = 0.

No reference has been made yet to the scaling with the small parameter *µ*. But by continuing to keep track of the scaling we would obtain the coefficients *h*_*ij*_ as power series of *µ*, hence

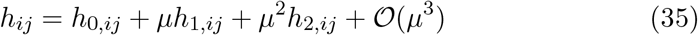

### 3.2 Block structure transformation with real transformation matrix

The analysis can be made easier by loosening the condition of exact diagonalizing of the Jacobian matrix to transformation into block structure, as e.g. excercised by [3, 4] in an SEIR epidemiological model for measles, going back to a first analysis in [5].

Observing the Jacobian matrix of the SIRUV around the endemic fixed point

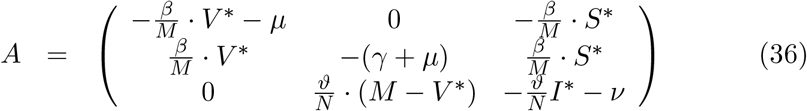

gives for *µ* → 0 the matrix

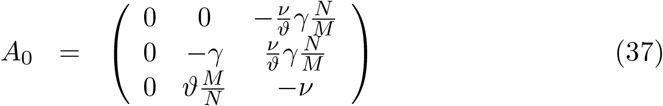

by remembering that *I*\* and *V* * are of order *µ* and 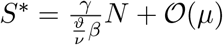

It will be useful later to dissect the matrix *A* = *A*_0_ + *A*_*r*_ into the zero order part *A*_0_ of the Jacobian matrix *A* and the rest *A*_*r*_ containing all higher orders of *µ* which can be easily done.

The eigenvalues of the matrix *A*_0_ are *λ*_1_ = 0, *λ*_2_ = 0 and *λ*_3_ = −(*γ* + *ν*), which corresponds to the zeroth order of the eigenvalues as we calculated in scaling before. Further, the eigenvectors of the lower right 2-dimensional square matrix are

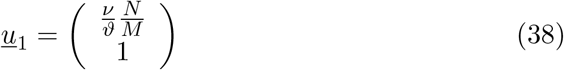

and

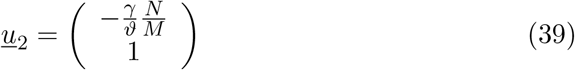

Hence a real transformation matrix 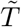 can be constructed via

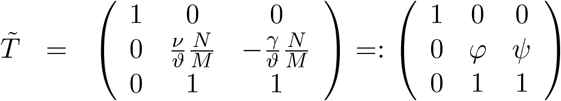

which has as inverse

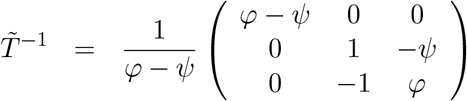

Then we have the transformation to block structure given by

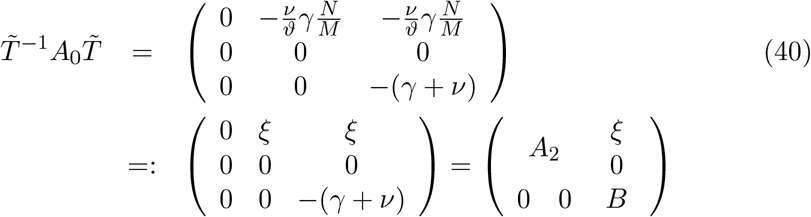

with

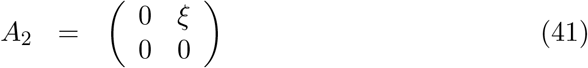

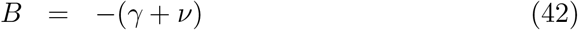

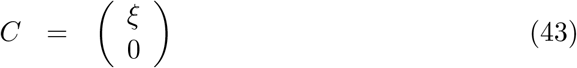

Now from

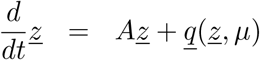

we have in zeroth order in *µ*

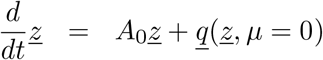

and with 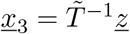 we obtain the block structure

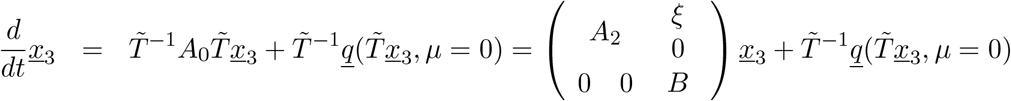

Hence in zeroth order in *µ* we have

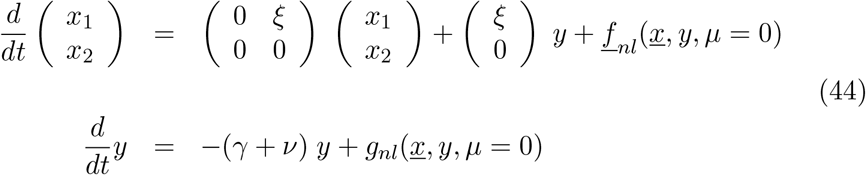

and with the blocks *A*_2_, *B* and *C*

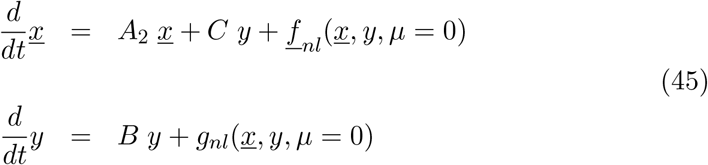

which has a structure for the center manifold analysis, see [9].

### 3.3 Families of center manifolds

Now for higher orders in *µ* we have to construct families of center manifolds [10] of the form

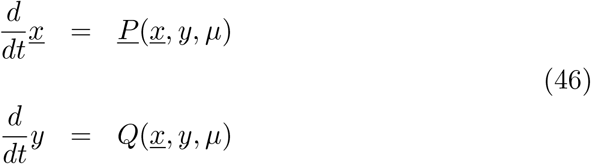

such that

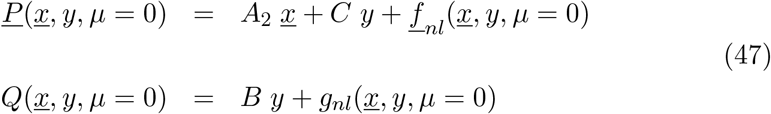

We first have to calculate the explicit form of the general *<u>P</u>* and *Q*, e.g. from Eq. (22)

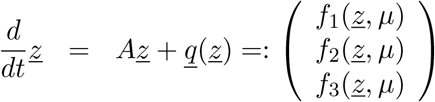

giving with 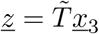 and 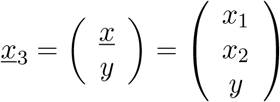 the expressions for *<u>P</u>* and *Q*

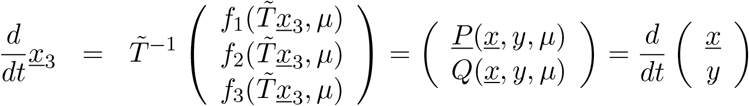

This gives us the families of center manifolds with parameter *µ*

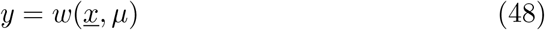

to be determined from

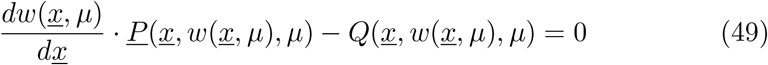

via series expansion

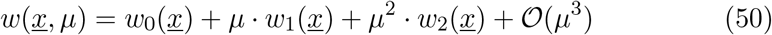

Especially we have for the center manifold in zeroth order *y* = *h*(*<u>x</u>*) from the equation system, Eq. (45),

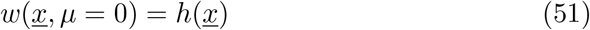

see [10].

### 3.4 Results for the SIRUV model

For the SIRUV model we have explicitly for

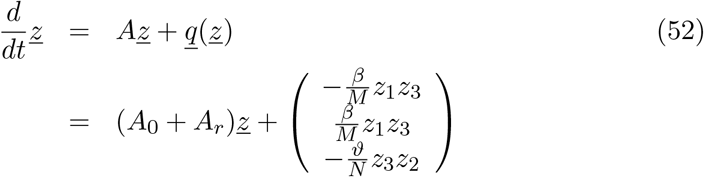

in which the Jacobian matrix *A* is decomposed into its zeroth order component *A*_0_ and the remaining *µ*-dependent rest *A*_*r*_ = *A*_*r*_(*µ*). Also it should be noted that the non-linear part *<u>q</u>*(*<u>z</u>*) is not dependent on *µ*. Explicitly, we have for *A* = *A*_0_ + *A*_*r*_ the result

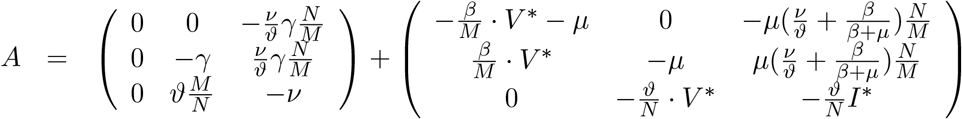

With the transformation matrix 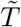 we then obtain

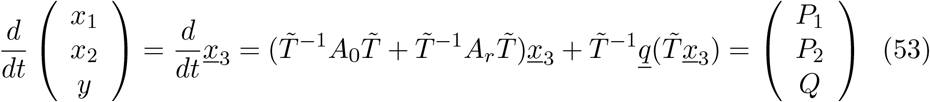

with the zeroth order terms

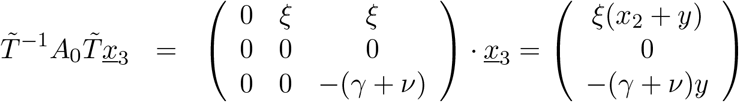

and

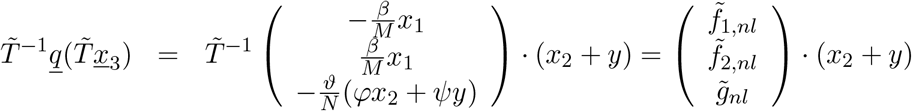

and the linear term of order *µ*

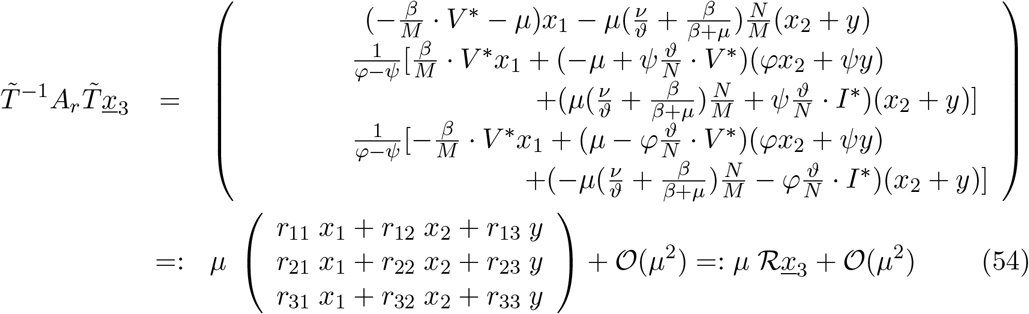

#### 3.4.1 Results for the SIRUV model in lowest order 𝒪 (*µ*^0^)

We have in zeroth order in *µ*

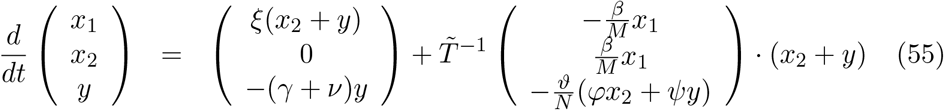

hence for *y* = *h*(*x*_1_, *x*_2_) we have the equation

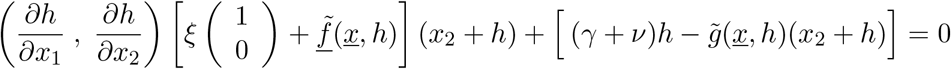

and with the ansatz in the leading quadratic terms

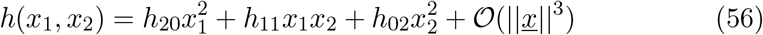

and after inserting and comparing the terms in 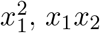 and 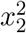 we obtain

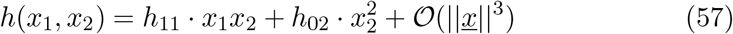

with 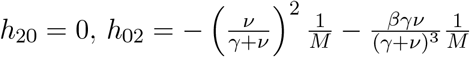 and 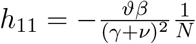

#### 3.4.2 Results for the SIRUV model in next to leading order *O*(*µ*)

In higher order in *µ* we have

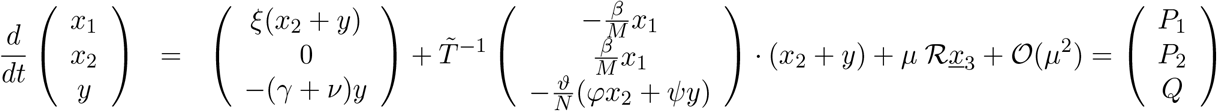

and hence for the family of center manifolds *y* = *w*(*<u>x</u>, µ*) we have

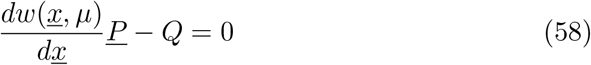

Further for *w* we have the ansatz

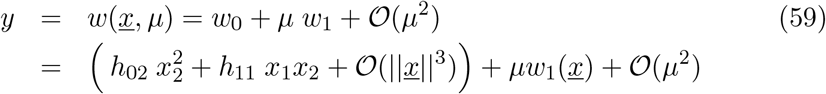

with *w*_0_(*<u>x</u>*) = *h*(*<u>x</u>*), and *h* as calculated before in zeroth order.

Now in approximation up to first order in *µ* with

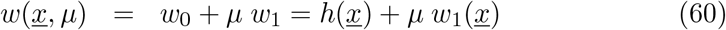

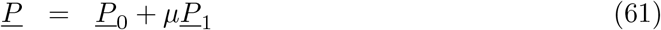

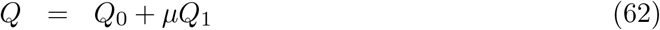

we have

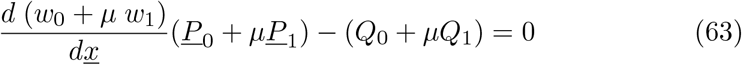

or

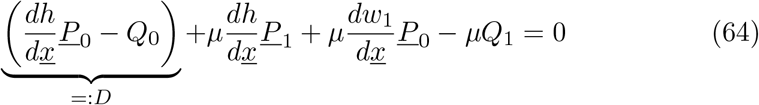

where we already know from the zeroth order calculations above that the leading term is 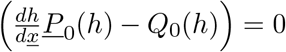 and that

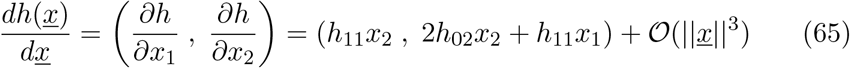

and finally

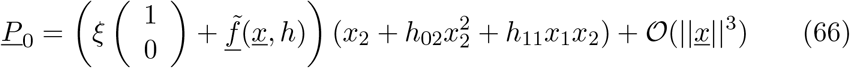

Further we have with *A* = *A*_0_ + *A*_*r*_ = *A*_0_ + *µA*_1_ + *µ*^2^*A*_2_ + 𝒪 (*µ*^3^)

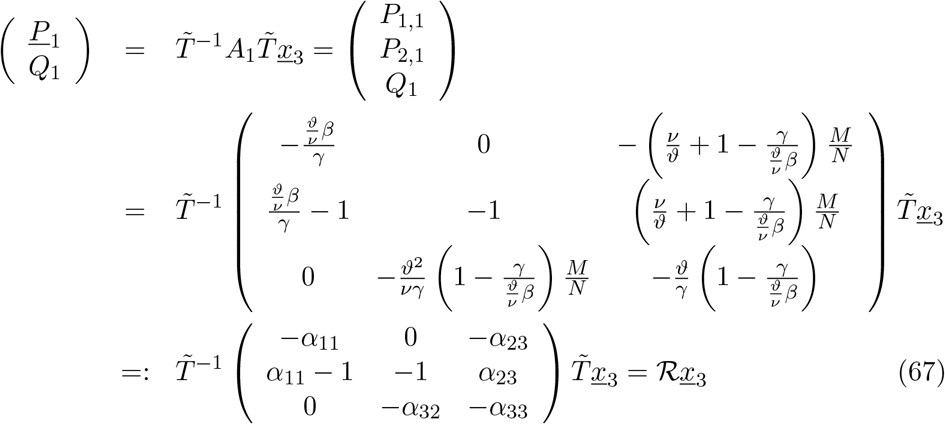

With this we have fully determined the terms in 𝒪 (*µ*) in

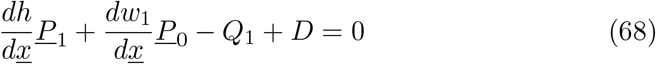

or in full as determining equation for *w*_1_(*<u>x</u>*) from

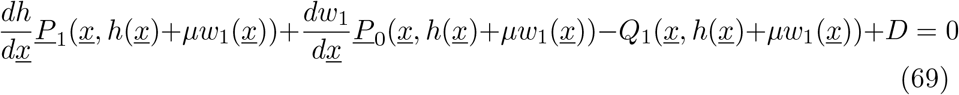

we simply have

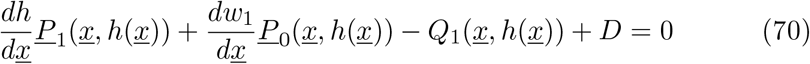

as next to leading order, since from Eq. (64) we see that Eq. (69) is already in first order in *µ*.

#### 3.4.3 Backtransformation to original coordinates and parameters in leading order

We have the center manifold in zeroth order as 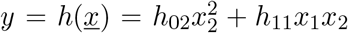. Closer to the original coordinates *z*_3_ = *V* (*t*) − *V*\* we have with the transformation 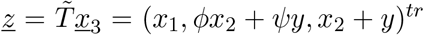, hence 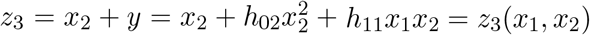, hence *z*_3_ = *V* (*t*) − *V* * as a function of *x*_2_. And from 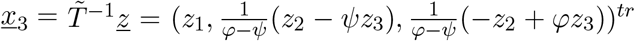, hence 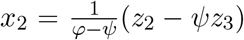 we can insert to obtain *z*_3_(*z*_2_), hence *z*_3_ = *V* (*t*) –*V\** as a function of *z*_2_ = *I*(*t*) –*I\**. Again cutting short, in lowest order we have 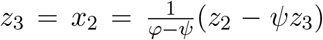 and hence

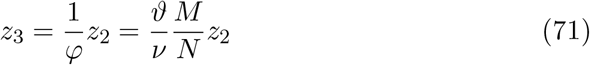

which gives

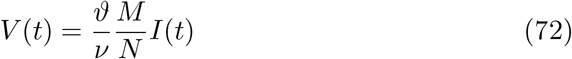

as final result. Eq. (72) hence gives in lowest order in *µ* the result *V* (*I*) proportional to *I*, which results in effective SIR-type models for the human infecteds only.

For the SISUV model in contrast, see Appendix B, we obtain with Eq. (147) in lowest order, now of *ε*, the Holling type II form for *V* (*I*), hence

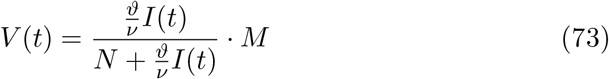

since in the SISUV model *I*\* and hence for long times *I*(*t*) is of order one and not of order *ε*. The simplification

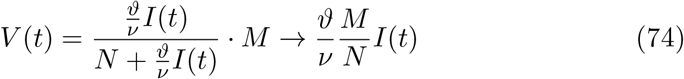

for biologically small parameter *µ* → 0 holds for the SIRUV model, if obtained by any method (e.g. as quasi-steady state assumption QSSA, see [2]), but not for the SISUV model. The present center manifold analysis of the SIRUV model gives in lowest order directly the simple linear relation.

## Data Availability

no empirical data used.

## 4 Acknowledgments

M. A. has received funding from the European Unions Horizon 2020 research and innovation programme under the Marie Skłodowska-Curie grant agreement No 792494. This research is also supported by the Basque Government through BERC 2018-2021 program and by Spanish Ministry of Sciences, Innovation and Universities: BCAM Severo Ochoa accreditation SEV-2017-0718. N.St. thanks the Mathematics Department of Trento University for the opportunity and support as guest lecturer in biomathematics, during which part of the present work has been conceived and started to be developed.

## A Eigenvalues of the Jacobian matrix around the endemic fixed point via Cardano’s method and its scaling

### A.1 Cardano’s method for finding the zeros of 3rd order polynomials

For any polynomial

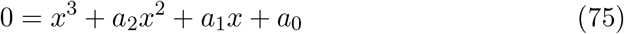

we can solve the equation by first reducing it via a coordinate transformation

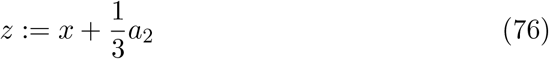

to the form

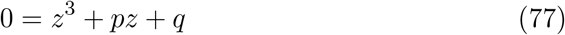

with

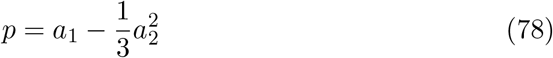

and

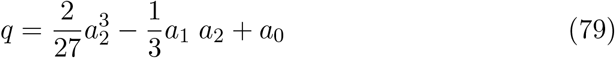

and further express *z* as sum of two variables *u* and *v*, hence

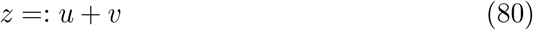

which gives the 3rd order polynomial in the form

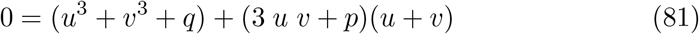

From this we get via setting the second bracket expression to zero the variable *v* as

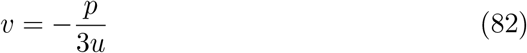

and setting th first bracket expression to zero we obtain for the variable *u*^3^ a solvable quadratic equation

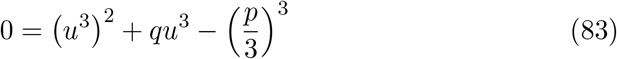

Hence we have

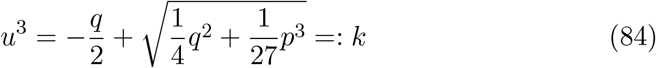

and 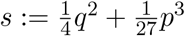.

Then the cubic Eq. (84) has three solutions with a phase factor 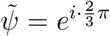, namely

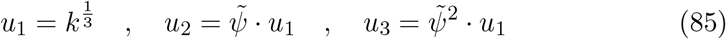

and their respective solutions *v*_*j*_ = −(*p/*3*u*_*j*_), hence *x*_*j*_ = *u*_*j*_ + *v*_*j*_ − *a*_2_*/*3 for *j* = 1, 2, 3. We have as powers of the phase factor 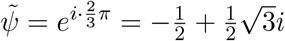 and 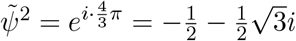, and of course 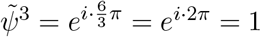.

### A.2 Application of Cardano’s method to the characteristic polynomial of the SIRUV Jacobian matrix

Implementing Cardano’s method for the parameters given in the SIRUV model and its Jacobian matrix we find

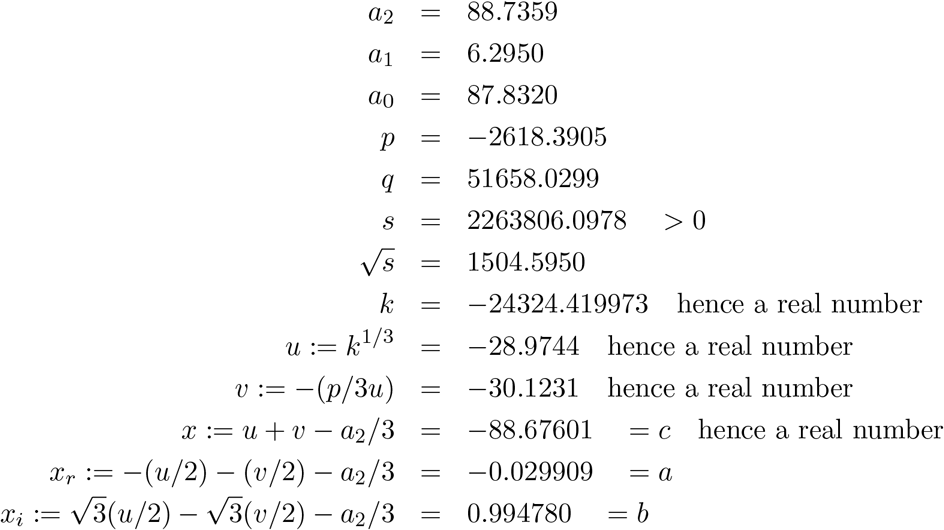

and hence gives the numerical values of the eigenvalues as follows

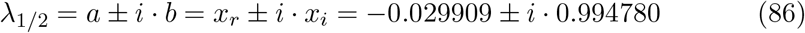

and

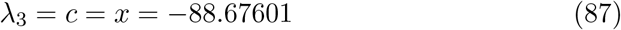

Now that we have the analytic form of the eigenvalues implicitly given via Cardano’s method, we can investigate the scaling of all relevant quantities with the small parameter *µ*. This will be done in the next section.

### A.3 Scaling with small parameter *µ*

First we observe numerically the scaling of all relevant quantities with the parameter *µ* by decreasing *µ* by a factor of 10 and comparing with the previous results from the original value of *µ*, hence

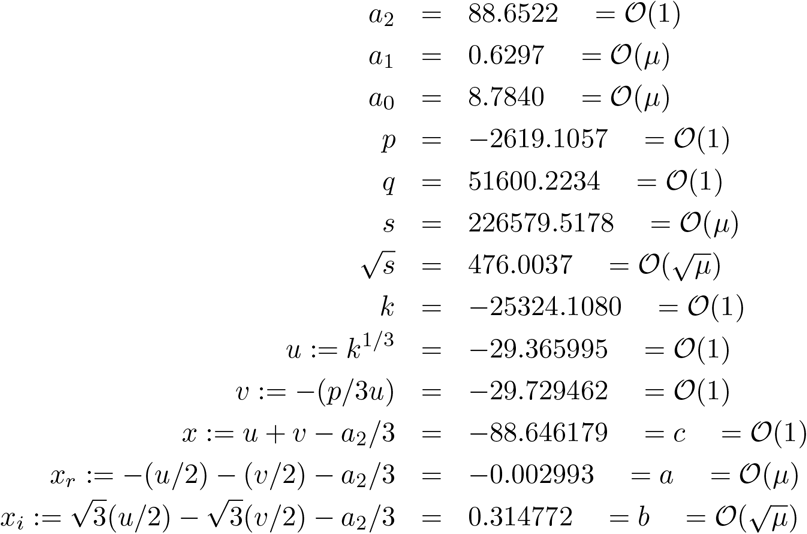

with many parameters of the Cardano method like *p, q, u* and *v* of or<u>d</u>er 𝒪 (1), w<u>h</u>ile the results *a* = − (*u/*2) − (*v/*2) *a /*3 = (*µ*) and 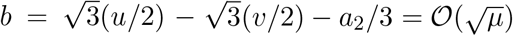 are of higher orders in *µ*.

Hence any scaling analysis has to take at least orders of *µ* into account, since order-one parameters in subtractions cancel the lowest order, and leave higher orders in *µ* only.

### A.4 Analytical results of the scaling analysis with small parameter *µ*

#### A.4.1 Scaling of endemic stationary state

For the stationary state we have the following scaling up to first order in *µ*

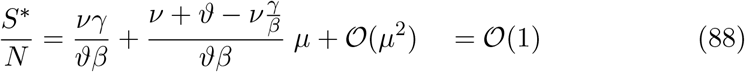

and

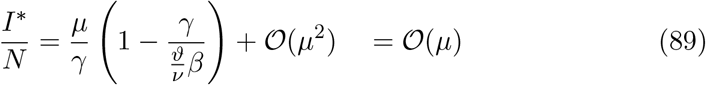

and

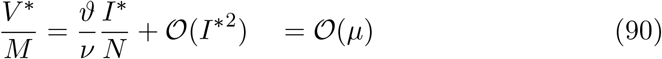

### A.4.2 Scaling of characteristic polynomial coefficients

The coefficients of the characteristic polynomial 0 = *λ*^3^ + *a*_2_*λ*^2^ + *a*_1_*λ* + *a*_0_ are in scaling up to first order in *µ* given by

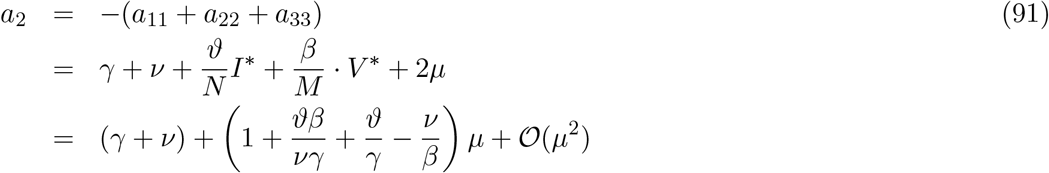

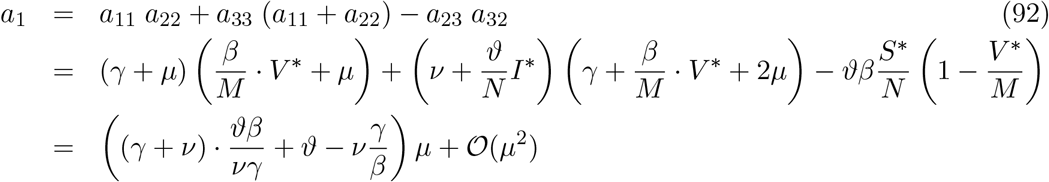

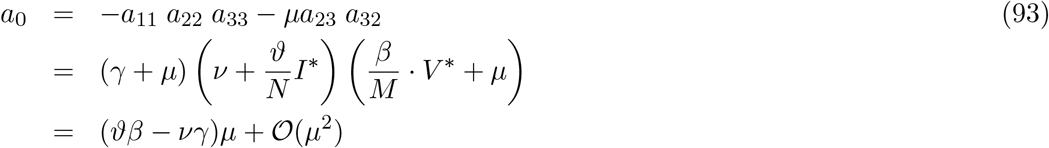

and are numerically tested.

#### A.4.3 Scaling of eigenvalues

For the scaling of the eigenvalues we need to take quantities like *c*_*µ*_ := *a*_2_ +*c* into account, where *a*_2_ = 𝒪 (1) and *c* = 𝒪 (1) but remembering that *a*_2_ is positive and *c* is negative we obtain *c*_*µ*_ = 𝒪 (*µ*). And for this we have to determine the order of *λ*_3_ = *c* from the whole Cardano procedure.

Once *c* is known, we can perform a polynomial divison of the characteristic polynomial 0 = *λ*^3^ + *a*_2_*λ*^2^ + *a*_1_*λ* + *a*_0_ obtaining

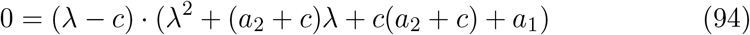

such that we then easily obtain the higher order quantities of the complex conjugated eigenvalues *λ*_1*/*2_ = *a ± i · b* as

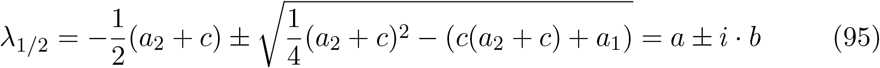

With the numerical values of *a*_2_ = 88.735880 and from Cardano’s method *c* = *x* = −88.676061 we obtain numerically *c*_*µ*_ = *a*_2_ + *c* = 0.059819, hence we have via the polynomial division

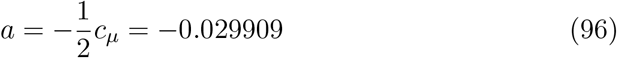

in complete agreement with the result from Cardano’s method. Further we can approximate *b* to order 𝒪 (*µ*^1*/*2^) as

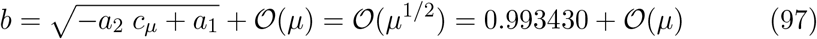

also in good agreement with the result from Cardano’s method of *b* = *x*_*i*_ = 0.994780. And scaling can be checked again by decreasing *µ* by a factor of 100, hence 𝒪(*µ*^1*/*2^) gives a factor of 10, with the result of *b* = *x*_*i*_ = 0.099546 and its approximation 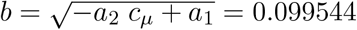.

The result of the scaling analysis for *c*_*µ*_ through out Cardano’s method is

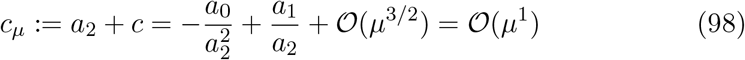

or with 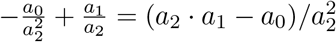 we have

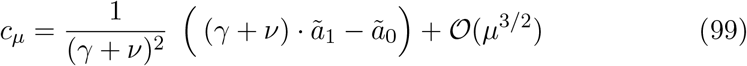

with *a*_1_ = *ã*_1_ + *O*(*µ*^2^) and *a*_0_ = *ã*_0_ + 𝒪(*µ*^2^), hence

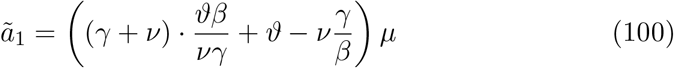

and

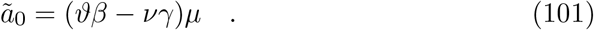

Or completely in terms of the original model parameters we have the final result of

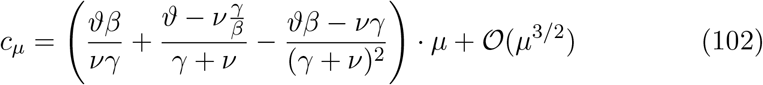

which gives the result of *c*_*µ*_ = 0.05981865 + 𝒪 (*µ*^3*/*2^) as compared to the exact result *c*_*µ*_ = *a*_2_ + *c* = 0.05986159.

Further we have the original 3rd eigenvalue given as

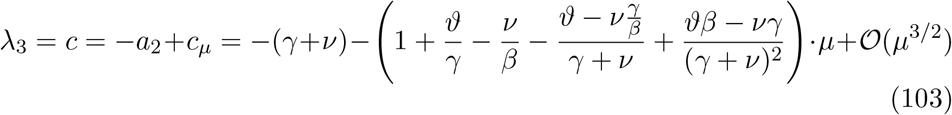

and the other two parameters of the first two eigenvalues *λ*_1*/*2_ = *a ± ib* are

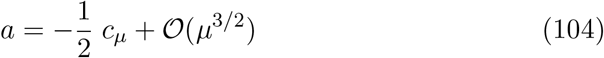

trivially and further

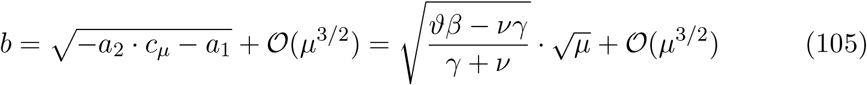

and numerically *b* = 0.995465 + 𝒪 (*µ*^3*/*2^) as compared to the exact result from Cardano’s method of *b* = 0.994780. And as check of the scaling when decreasing *µ* again by a factor of 100, we obtain as approximation *b* = 0.09954648+𝒪 (*µ*^3*/*2^) as compared to the exact *b* = 0.09954580.

## B Center manifold analysis for the SISUV model with scaling analysis

The SISUV model is given by

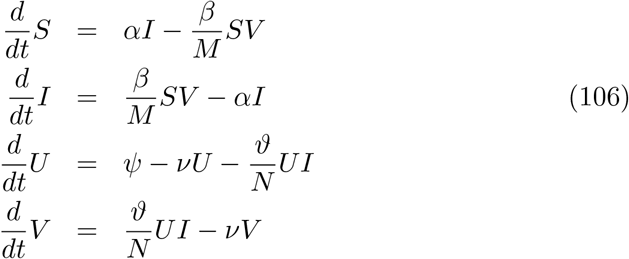

with all parameters as also used in the SIRUV model, but 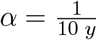 and *β* = 2*·α*, matching qualitatively the original SIRUV model, just with slow parameters *α* and *β* for the human dynamics. The two dimensional dynamical system then, by taking the conservations for human population *N* = *S* +*I* and for mosquitoes *M* = *U* + *V* into account, is given as

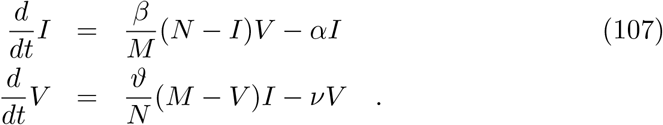

The time scale separation is now given by the rescaling of the slow parameters 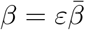 and 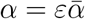. The endemic stationary state is given by

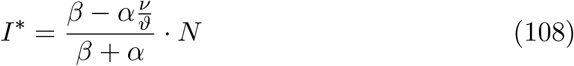

and as before

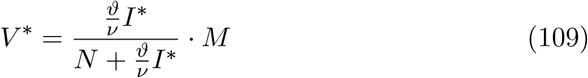

### B.1 Linear part of the center manifold analysis for the SISUV model

Then the Jacobian matrix around the endemic stationary state is given by

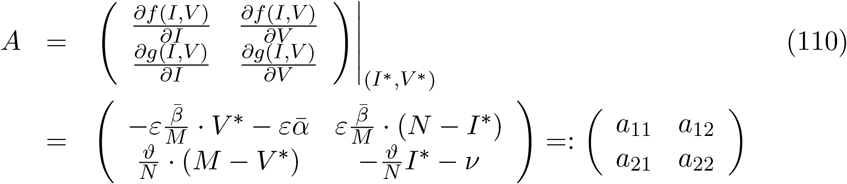

or in scaling with *ε*

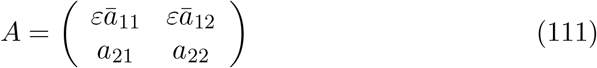

The eigenvalues are given by

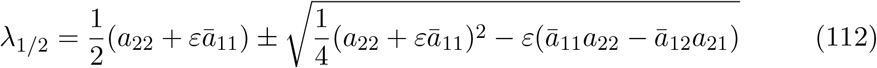

hence in leading order 𝒪 (*ε*^0^) we have

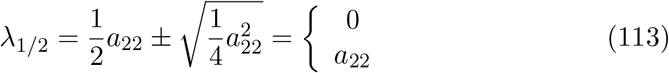

since *a*_22_ is negative and hence 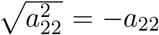. This result was used previously in the center manifold analysis of the SISUV model [1]. Now in next to leading order we have the following result, by using

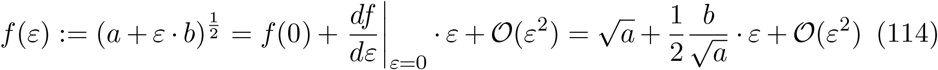

giving

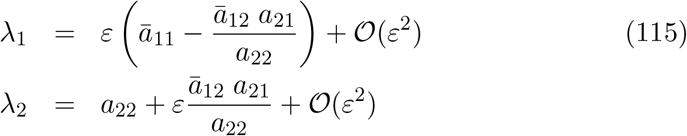

hence

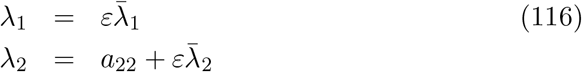

with 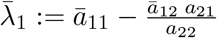 and 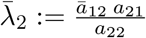.

Then the eigenvectors can be given as

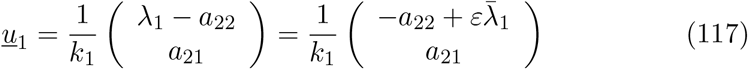

with eventually normalization constant 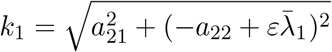 and

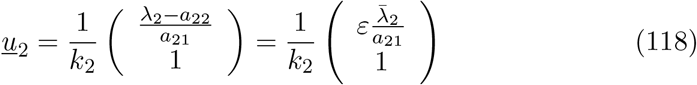

with eventually normalization constant 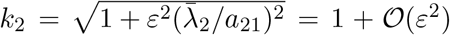, see footnote^1^. And with this we obtain the transformation matrix *T* and its inverse *T*^−1^ via *AT* = *T* Λ with *A* the Jacobian matrix as given above, and the matrix Λ the diagonal matrix of eigenvalues, with

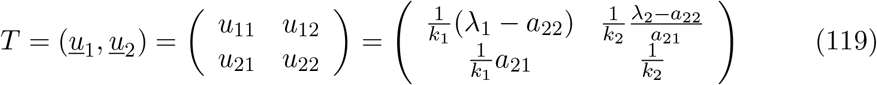

or in scaling with *ε*

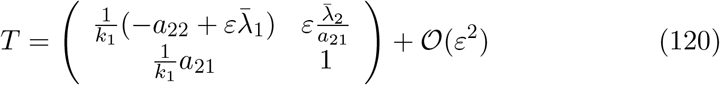

(where the terms proportional to 1*/k*_1_ could be analyzed further in scaling, but eventually disturbing the normalization and the geometric interpretation, this could be checked later in the calculations). The inverse *T*^−1^ is given by

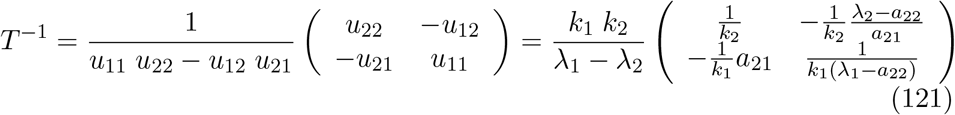

and in scaling

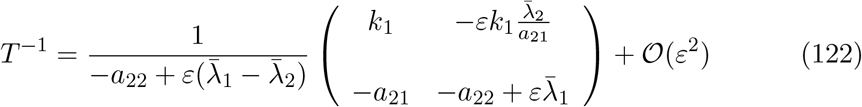

### B.2 Nonlinear part of the center manifold analysis for the SISUV model

With the distance of the dynamical system from the endemic fixed point (*I*\*, *V* *) given by

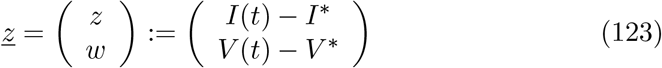

we have the linearized dynamics via the Jacobian matrix *A* given as

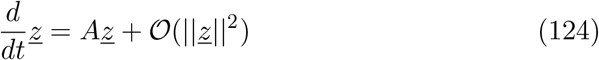

and with the eigenvalue/eigenvector analysis *AT* = *T* Λ the diagonalization of the dynamics

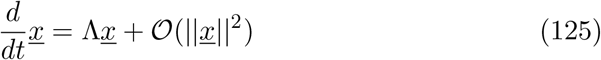

with the transformation

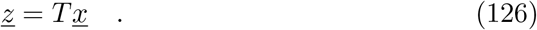

Now we take the nonlinear part of the dynamics <u>*q*</u> explicitly into account via

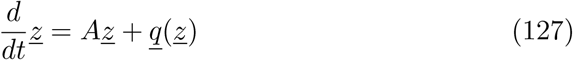

with

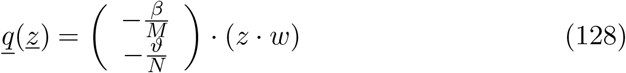

and with the transformation *<u>z</u>* = *T<u>x</u>* for *<u>x</u>* =: (*x,y*)^*tr*^ we obtain

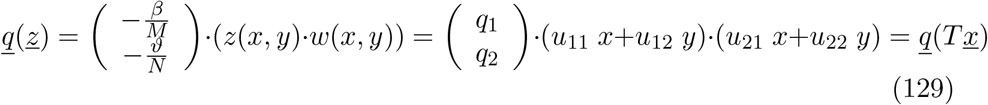

hence

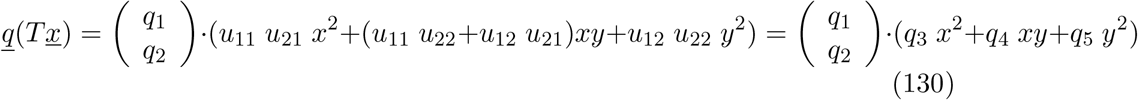

with constants 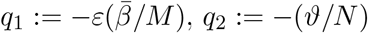 and *q*_3_, *q*_4_ and *q*_5_ given via

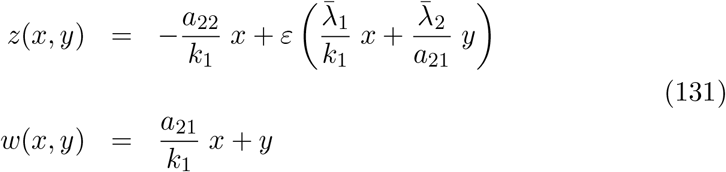

such that we have now the complete nonlinear dynamical system in transformed variables for the diagonalized linear part as

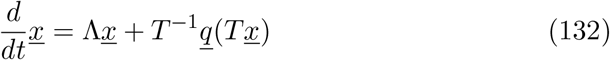

with the nonlinear part given by

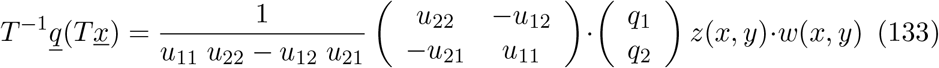

and abbreviating

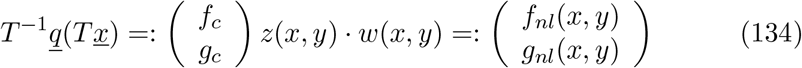

with constants

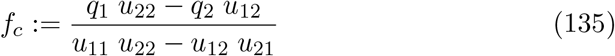

and

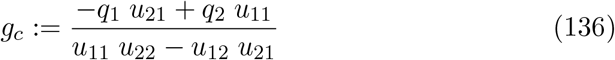

giving explicitly and in scaling

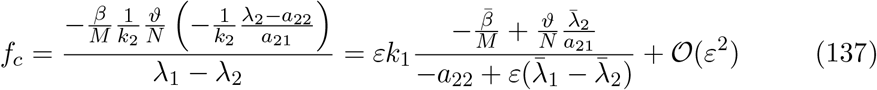

and

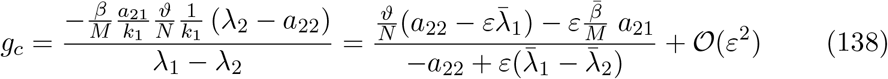

Hence we have the dynamical system now as

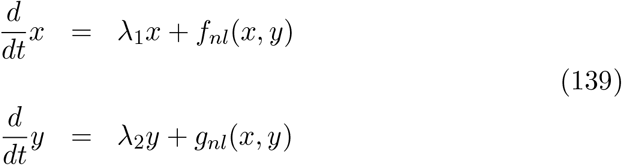

and assuming the fast variable *y* as a function of the slow variable *x*, hence *y* = *h*(*x*), we have by using the chain rule of differentiation

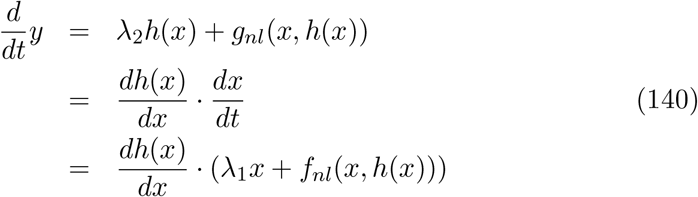

This gives the functional *𝒩* (*h*(*x*)) of the unknown function *h*(*x*) as a functional equation *𝒩* (*h*(*x*)) = 0 to be solved with

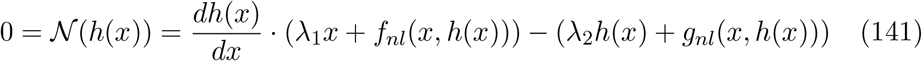

This could be done with a polynomial ansatz for *h*(*x*) := *h*_2_*x*^2^ + *h*_3_*x*^3^ + *h*_4_*x*^4^ + 𝒪 (*x*^5^) around the center, hence small *x* [1], giving recursions for higher orders of *h*_*ν*_ as functions of lower, similar to what is reported in [8].

But given the analysis of all terms in scaling with the small parameter *ε*, we find that as well *λ*_1_ = 𝒪 (*ε*) as *f*_*nl*_(*x, h*(*x*)) = 𝒪 (*ε*) due to *f*_*c*_ = 𝒪 (*ε*). Hence in leading order 𝒪 (1) we have from Eq. (141) only an algebraic equation left

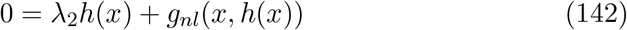

So we can in this case use the same technique as in singular perturbation, namely we expand *h* in powers of the small scaling parameter *ε* with

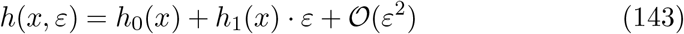

such that Eq. (141) can be solved successively in increasing orders of *ε*, starting with lowest order

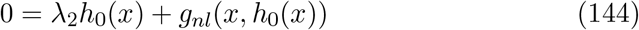

which can be solved directly giving

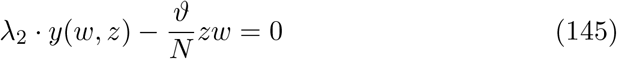

or

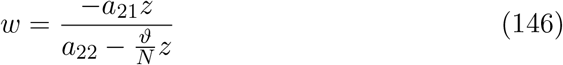

and with *z* = *V* (*t*) − *V* * and *w* = *I*(*t*) − *I*\* we obtain

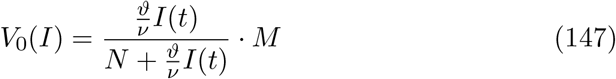

analogously to the result from singular perturbation via the ansatz *V* (*I, ε*) = *V*_0_(*I*) + *V*_1_(*I*) *· ε* + 𝒪 (*ε*^2^) directly applied to Eq. (107) and with the scaling 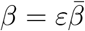 and 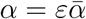.

And then as in singular perturbation the next order terms can be calculated using the lowest order result to calculate the next order via Eq. (141). In this respect center manifold analysis gives the same result to all orders as singular perturbation in the cases where 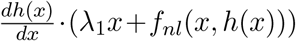 is of higher order than *λ*_2_*h*(*x*) + *g*_*nl*_(*x, h*(*x*)), i.e. the standard time scale separation form *dx/dt* = *εf* and *dy/dt* = *g*.

Just in cases not in standard time scale separation form, the Eq. (141) can be solved via *h* expanding in small scaling parameter *ε* and in slow state variable *x*, and then goes beyond singular perturbation (as we see in the SIRUV system in the main text). Else, singular perturbation gives already the answer of the slow mainfold *y* = *h*(*x, ε*), and center manifold analysis only adds the linear algebra part of eigenvalues and eigendirections.

## C Transition from slow-fast time scale separation to final spiraling on center manifold

From the scaling of the state variables at the endemic fixed point, Eqs. (88), (89) and (90), we have *I*\* = 𝒪 (*µ*) and with it also *V* * = 𝒪 (*µ*), whereas *S*\* = 𝒪 (1). We can apply this as scaling transformations for the state variables outside the endemic equilibrium, hence

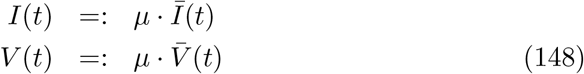

with *Ī* (*t*) and 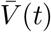 in roughly the magnitude of *S*\*. By inserting this transformation into the dynamic equations, Eq. (2), we obtain for the variables *S*(*t*), *Ī* (*t*) and 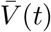 the dynamical system

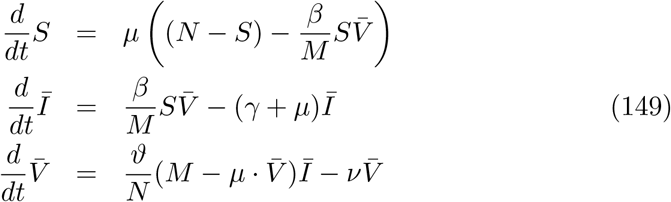

which has now in the small parameter *µ* one slow variable, namely *S*, and two fast variables, 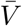 and *Ī*.

In this form with variables *S*, 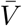 and *Ī* we have exactly the standard singular perturbation form given by

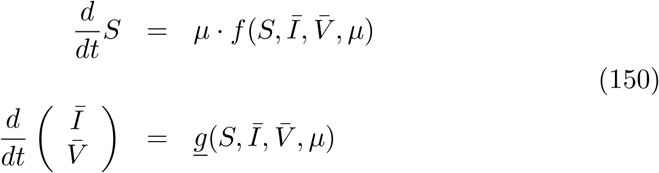

with one slow and two fast variable. This, of course, cannot be the dynamic regime close to the endemic fixed point, since there we have two slow variables *S* and *I* and one fast variable *V*, but rather describes a time scale separable regime in the transient phase, where we observe from a state of low numbers of susceptibles a slow building-up phase of susceptibles *S* with very low numbers of infected humans *I* and infected mosquitoes *V*, hence *I*\* = 𝒪 (*µ*) and *V* * = 𝒪 (*µ*), and then in a state with many susceptibles a fast burst of infecteds, infected humans *I* and infected mosquitoes *V*, until they burn out more susceptibles than would be needed to keep a high infection level. This fast bursting phase is then followed again by a slow rebuilding of susceptibles and again in low numbers of infected, *I* and *V*.

The slow-fast build-up and burst dynamics can be observed in the SIRUV model, as it can be in other SIR-type models (see [12] for such slow-fast dynamics in some SIR-type models), when starting the dynamics with initial conditions far away from the endemic fixed point, see Fig. 3. In a) we observe from initially very low numbers of infected a sudden spike of number of infected (just before *t* = 20), followed by another phase of very low numbers of infected, in which the susceptibles are built up again, followed by another spiking. This is the slow-fast dynamics phase. However, gradually the spikes decrease and the troughs in the low infection phases increase, until the dynamics finally enters into the oscillatory phase around the endemic fixed point, the regime which is described by the time scale separation towards the center manifold. The relation between bursting infected and susceptibles re-building becomes more obvious in the state space plot in Fig. 3 b), where from about 160 000 susceptibles and very low numbers of infected the susceptibles reach a level of about 340 000, from which on the number of infected rapidly increases, before crashing again at numbers of susceptibles around 180 000 etc. Finally, we can also observe the spiraling into the endemic fixed point at around 250 000 susceptibles, similar to what was observe in Fig. 1 b), there with initial conditions close to the endemic fixed point.

**Figure 3:**
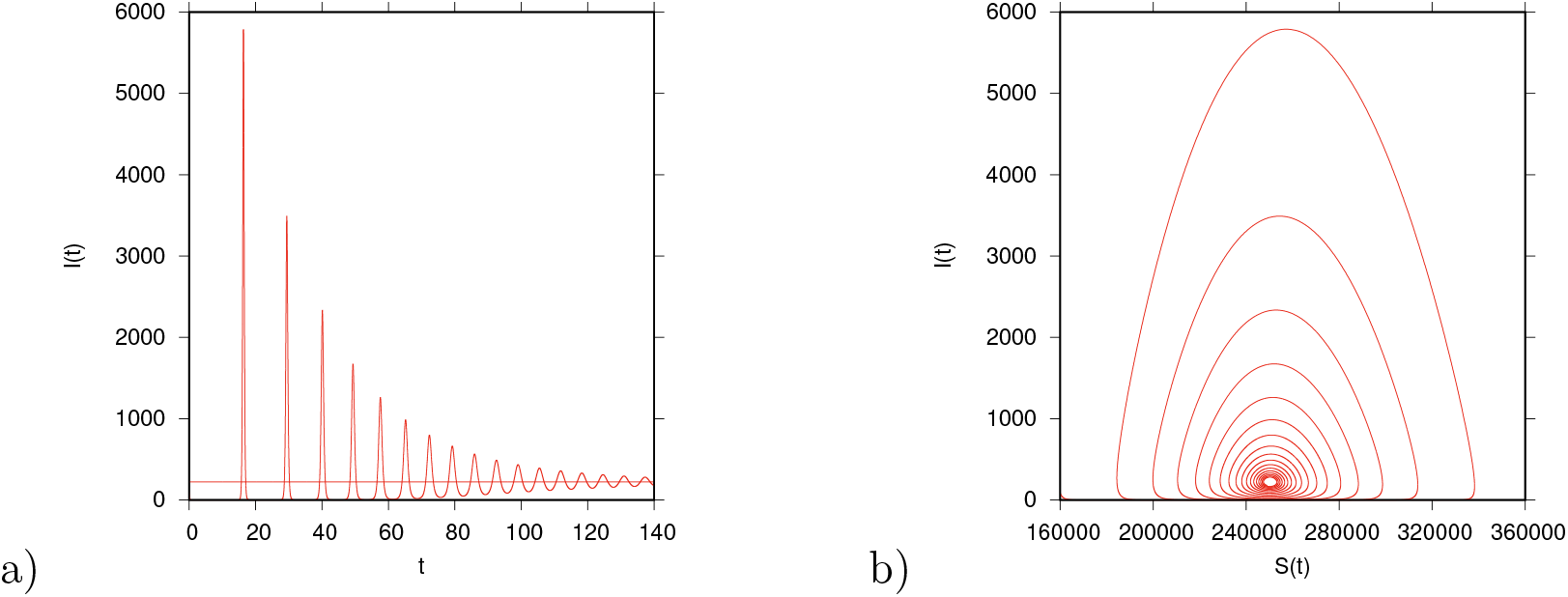
Time series of infected and state space plot of infected versus susceptibles in the SIRUV model. Initial conditions far away from the endemic fixed point, S_0_ = 0.17 · N, I_0_ = 0.0001 · N, and small number or no infected mosquitoes. After long times with low infection rate and slow build up of susceptibles we observe fast large outbreaks of infected, burning out large proportions of susceptibles (slow-fast regime). Then we observe a gradual transition to oscillations into the endemic fixed point with the known dynamics in the vicinity of the fixed point described (center manifold regime).

## D Quadratic approximation of center manifold

From

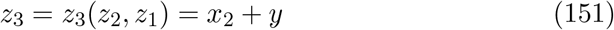

and

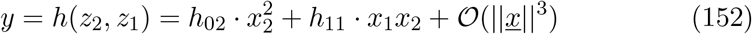

we obtain with the transformations from the center manifold coordinates *<u>x</u>* to the original coordinates *<u>z</u>*, remembering that *z*_1_ := *S*(*t*) − *S*\*, *z*_2_ := *I*(*t*) − *I*^*^ and *z*_3_ := *V* (*t*) − *V* *, with

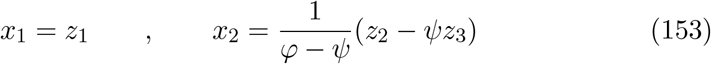

the expression

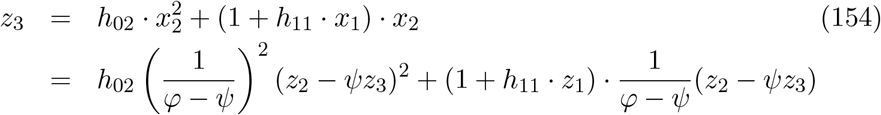

which has to be solved for *z*_3_ = *z*_3_(*z*_2_, *z*_1_).

The quadratic equation for *z*_3_ is given by

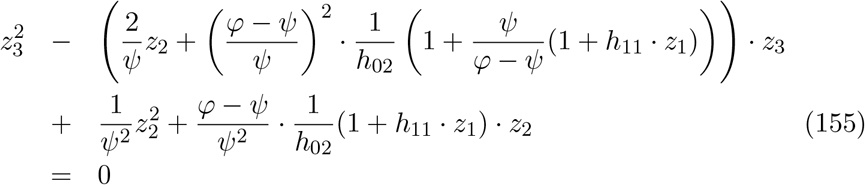

hence with

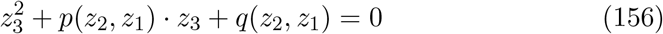

the solution *z*_3_(*z*_2_, *z*_1_) is given by

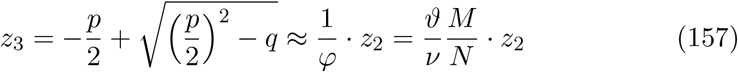

and plotted graphically for *z*_1_ = 0 in Fig. 4. The coefficients for the quadratic approximation of the center manifold *z* = *h*(*x*_1_, *x*_2_) are 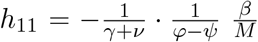, giving in original model parameters 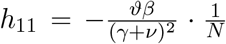, and furthermore the second coefficient 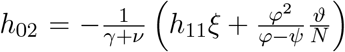, which is in original model parameters 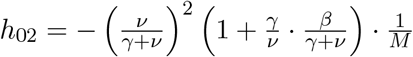.

**Figure 4:**
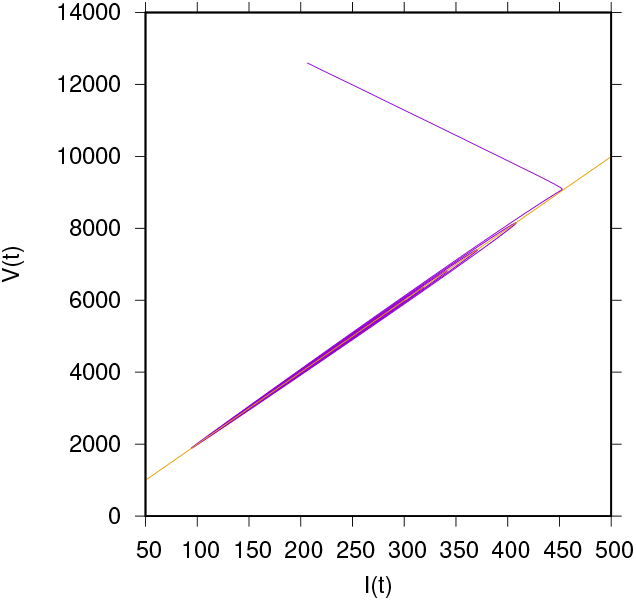
Quadratic approximation of z_3_ = z_3_(z_2_, z_1_) for continuously varying z_2_ and z_1_ = 0 in comparison with the trajectory spiralling into the fixed point. The quadratic expression gives essentially the linear relation between V and I again, namely 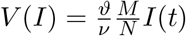.

In Fig. 5 we explore the *z*_1_ dependence of *z*_3_ = *z*_3_(*z*_2_, *z*_1_) further by comparing the graphs for *z*_1_ = 242000 − *S*\*, the smallest value of *S* when spiralling into the fixed point and *z*_1_ = 258000 −*S*\*, the largest value of *S* when spiralling into the fixed point. The slight variations of the lines for the varying *z*_1_ values around the value *z*_1_ = 0, the mid point, see. Fig. 4, are in the range of the variations of the oscillating trajectory.

**Figure 5:**
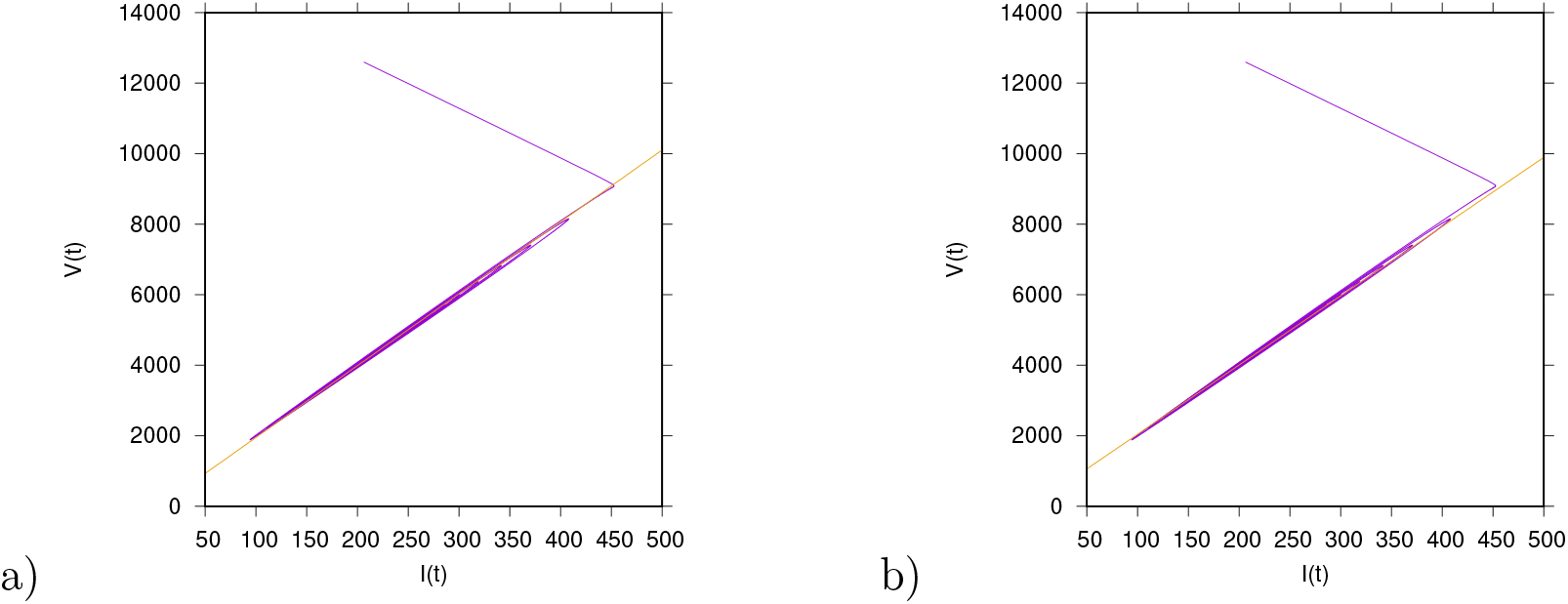
Quadratic approximation of z_3_ = z_3_(z_2_, z_1_) for continuously varying z_2_ and different values of z_1_, a) z_1_ = 242000 − S* and b) z_1_ = 258000 − S*, values in the range of the oscillations of S(t) into the fixed point.

## E Quadratic approximation of center manifold in next to leading order in *µ*

### E.1 Analysis of determining equation of *w*_1_

For the family of center manifolds

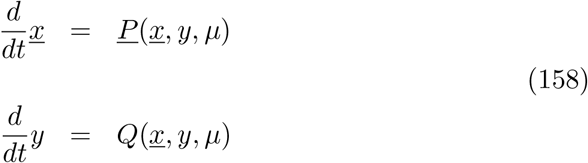

we have for *y* = *w*(*<u>x</u>, µ*) the detrmining equation for *w*(*<u>x</u>, µ*) given as

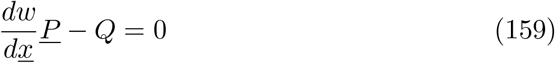

and expand in orders of *µ* in the following way

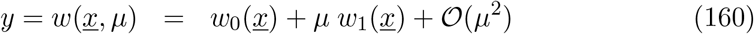

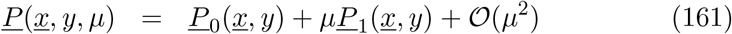

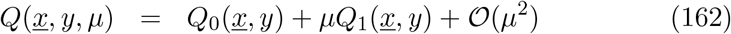

taking care of all variable-dependences, and remembering from zeroth order in *µ* that *w*_0_(*<u>x</u>*) = *h*(*<u>x</u>*). Hence in next to leading order in *µ* we have

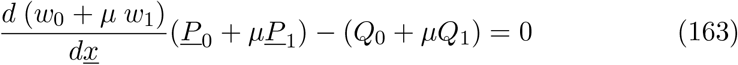

or

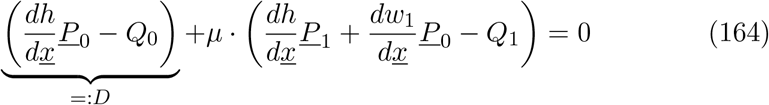

where we still have to evaluate *<u>P</u>*_*i*_(*x, y*) and *Q*_*i*_(*<u>x</u>, y*) with *y* = *h*(*<u>x</u>*) + *µ w*_1_(*<u>x</u>*). It is explicitly

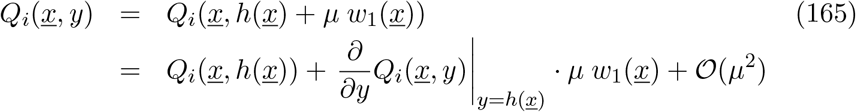

and respectively for *<u>P</u>*_*i*_(*<u>x</u>, y*). This gives for 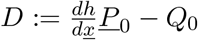 the result to first order in *µ* as

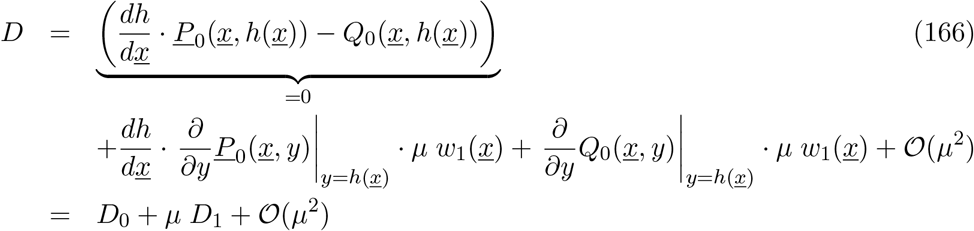

with *D*_0_ = 0 from the zeroth order calculation in *µ* by determining *h*(*<u>x</u>*). Further considering terms like

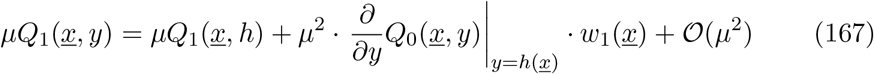

we obtain in 𝒪 (*µ*) the determining equation for *w*_1_(*x*) as

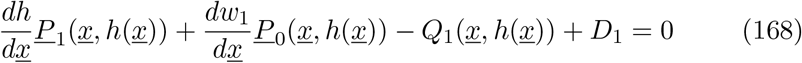

with

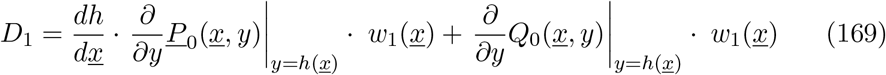

and hence finally

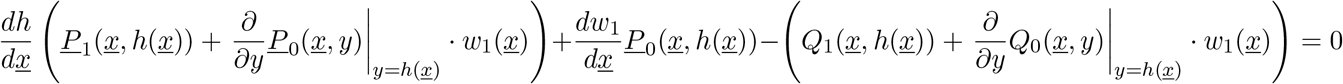

taking care of all variable dependences, or in short hand notation

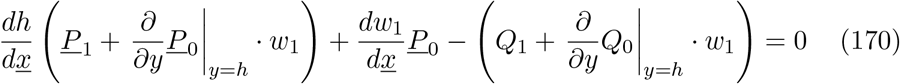

From this equation we now can calculate *w*_1_(*<u>x</u>*) in quadratic order, since we already have *h*(*<u>x</u>*) = *w*_0_(*<u>x</u>*) in this quadratic order, with the ansatz for *w*_1_(*<u>x</u>*) in the form

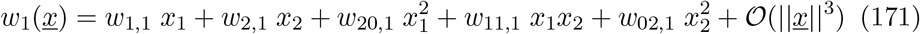

### E.2 Calculation of functions *<u>P</u>* _0_, *Q*_0_, *<u>P</u>* _1_, etc

We now calculate the functions *<u>P</u>* _0_(*<u>x</u>, y*), *Q*_0_(*<u>x</u>, y*), *<u>P</u>* _1_(*<u>x</u>, y*), *Q*_1_(*<u>x</u>, y*) and the derivatives 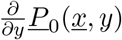 and 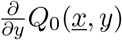 to order 𝒪 (||*<u>x</u>*||^2^), remembering that *y* = *h*(*<u>x</u>*) to be inserted is of quadratic order, in a form that we can then easily access zeroth order, linear and quadratic terms in *<u>x</u>*.

First we calculate *<u>P</u>*_0_ and *Q*_0_ such that we then can easily take the derivatives in respect to *y*. We have from

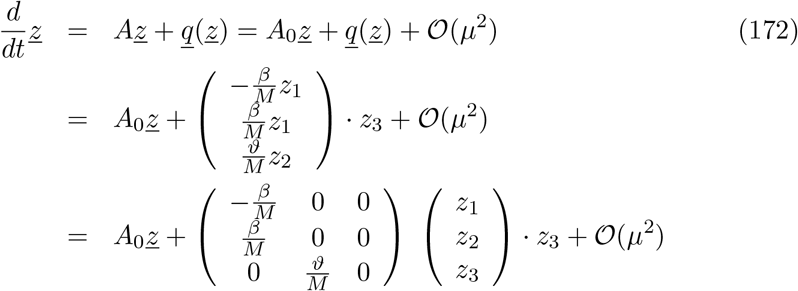

with the matrix

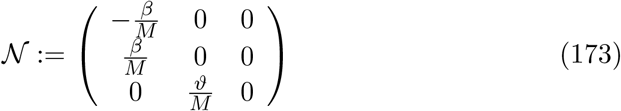

in coordinates 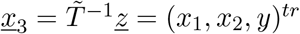

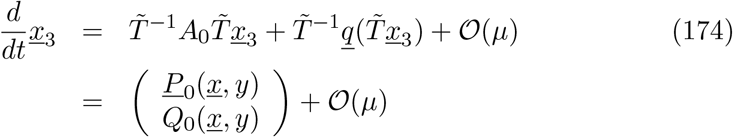

with *<u>x</u>* = (*x*_1_, *x*_2_)^*tr*^.

Hence with the explicit transformation *z*_3_ = *x*_2_ + *y* we obtain the functions *<u>P</u>* _0_(*<u>x</u>, y*) and *Q*_0_(*<u>x</u>, y*) via

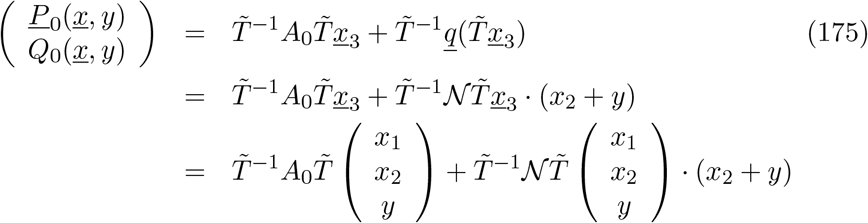

with the matrices

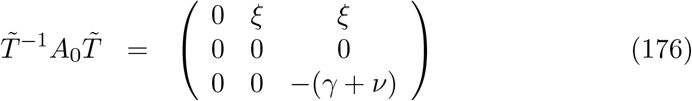

and

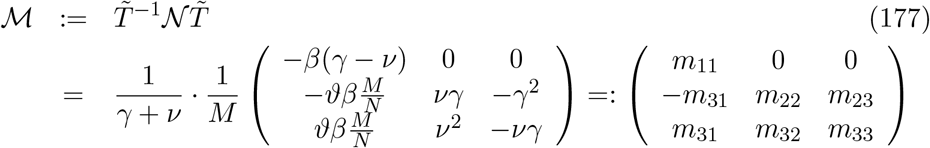

Hence we obtain for *<u>P</u>* _0_(*<u>x</u>, y*) explicitly

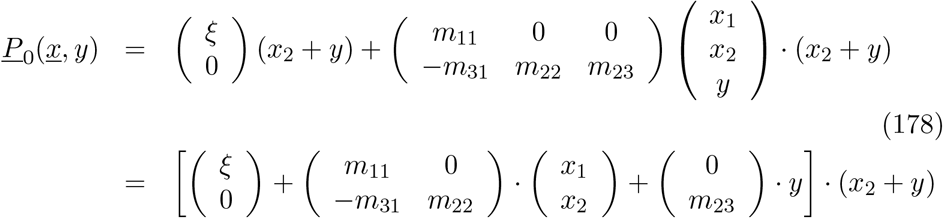

and for *Q*_0_(*<u>x</u>, y*) explicitly

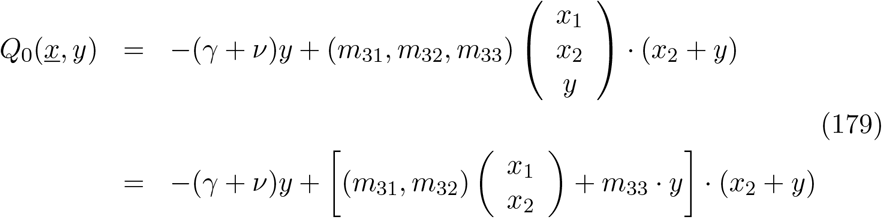

and from these also the derivatives

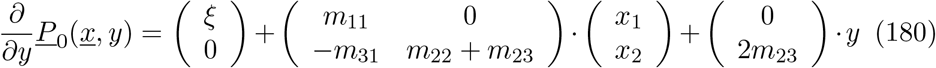

and

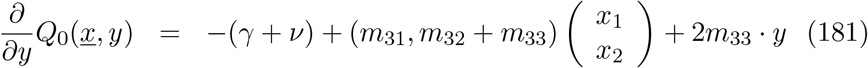

To determin *<u>P</u>* _1_ and *Q*_1_ we have to take the expansion of *A* = *A*_0_ +*µA*_1_ + 𝒪 (*µ*^2^) into account, since the nonlinear part *q*(*<u>z</u>*) is *µ*-independent in

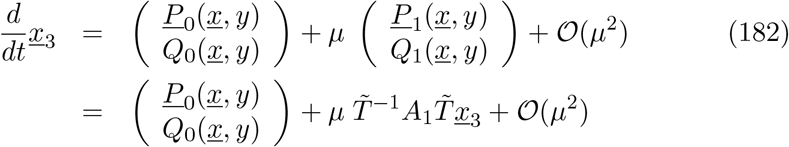

and hence

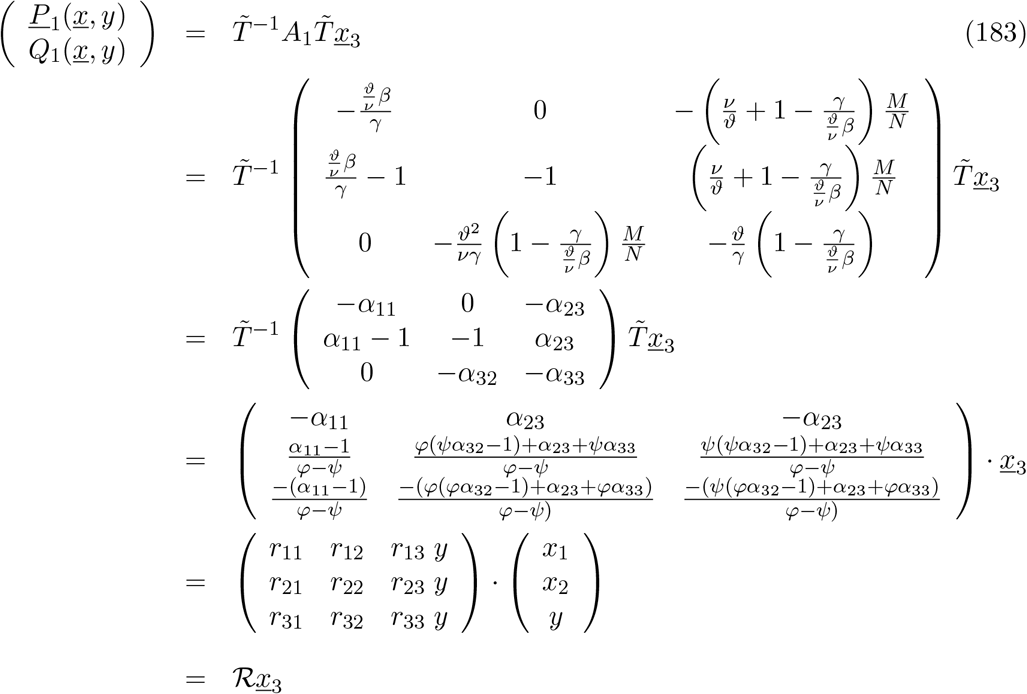

such that we obtain *<u>P</u>* _1_ and *Q*_1_ as

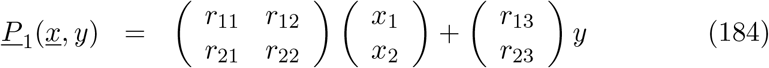

and

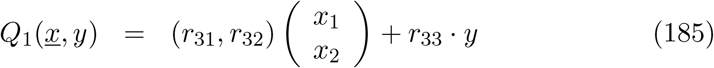

with *r*_*ij*_ the matrix entries of ℛ.

### E.3 Calculation of coefficients of *w*_1_ to second order in *<u>x</u>*

With the functions *<u>P</u>* _0_, *Q*_0_, *<u>P</u>* _1_, *Q*_1_ and 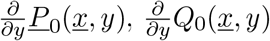 calculated as

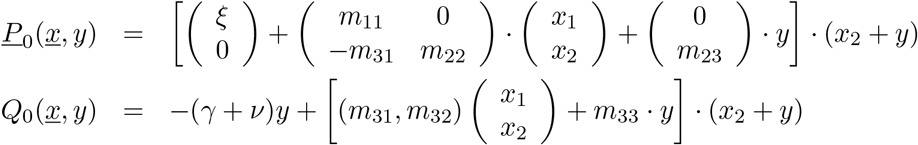

and from order 𝒪 (*µ*)

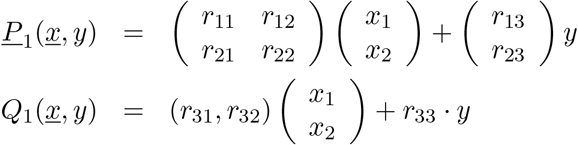

and the derivatives

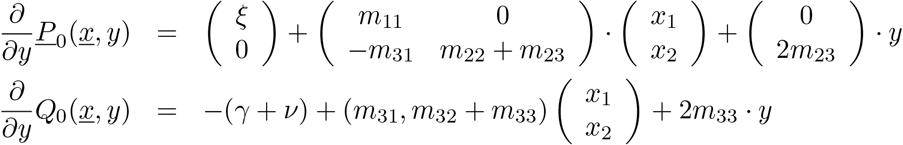

we can now evaluate the *w*_1_(*<u>x</u>*)-determining equation, Eq. (170), by inserting *y* = *h*(*<u>x</u>*) = 𝒪 (||*<u>x</u>*||^2^), with

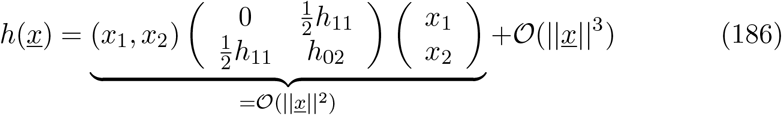

and its derivative

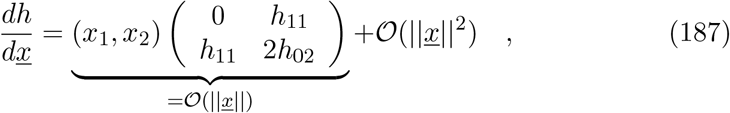

into all functions *<u>P</u>* _0_, *Q*_0_, *<u>P</u>* _1_, *Q*_1_ and 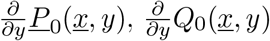 and determin the orders in *<u>x</u>* of the terms like

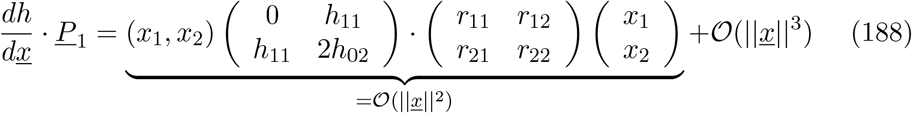

with the following results for the five terms involved in Eq. (170):

#### E.3.1 Term 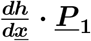

From Eq. (188) we obtain the term 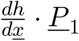

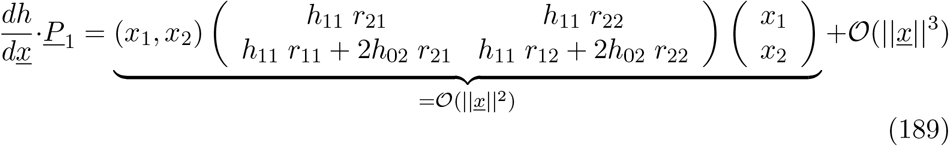

as one quadratic form plus higher order terms in *<u>x</u>*. In this term no linear part is appearing.

#### E.3.2 Term 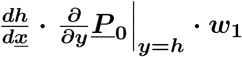

For the second term in Eq. (170) we have

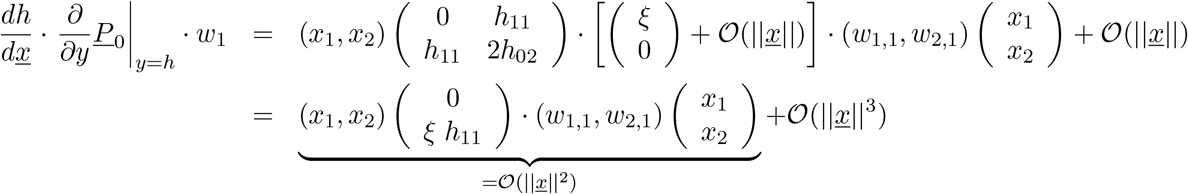

with the quadratic matrix

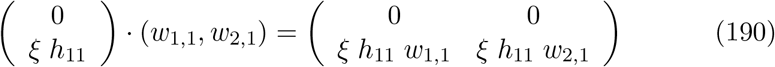

for the quadratic form of the term 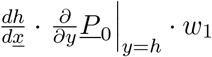.

#### E.3.3 Term 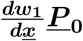

For the third term we have

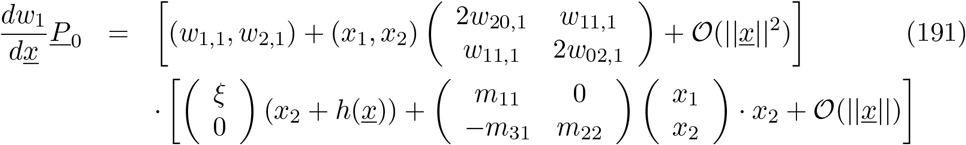

and after some calculation, and baring in mind that only

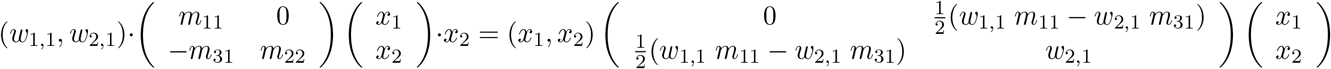

gives a standart quadratic form again, giving finally

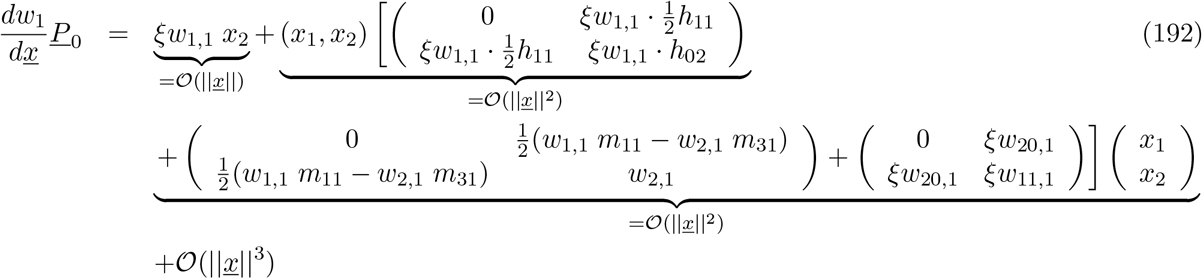

with two more terms of Eq. (170) to be evaluated.

#### E.3.4 Term *Q*_1_

The term

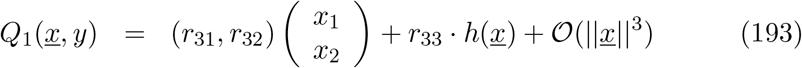

giving

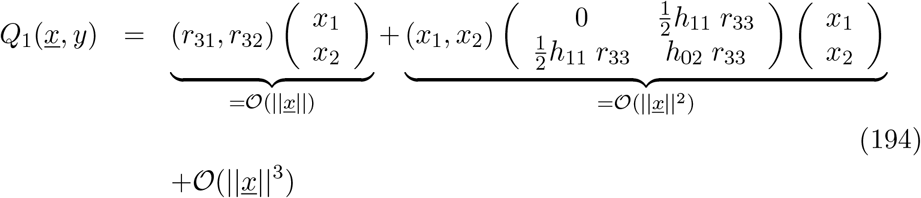

has a linear part, not involving the coefficients *w*_1,1_ and *w*_2,1_, hence will finally give rise to non-vanishing linear coefficients in *w*_1_(*<u>x</u>*).

#### E.3.5 Term 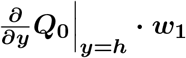

Finally, we evaluate the term

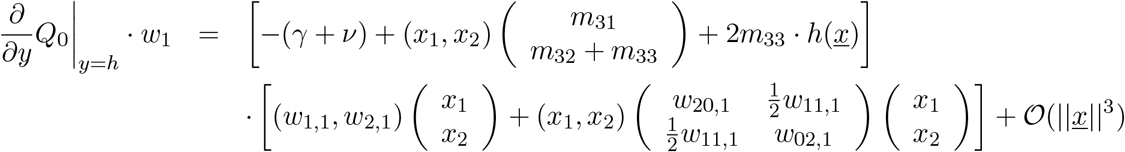

giving

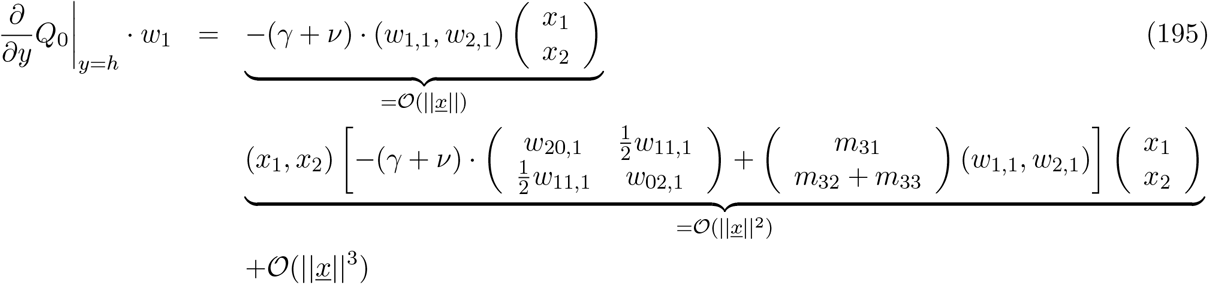

so that now we have all terms of Eq. (170) and can hence calculate the coefficients of *w*(*<u>x</u>*).

#### E.3.6 Calculaton of the linear coefficients of *w*_1_

We now collect all linear terms in *<u>x</u>* in Eq. (170) to determin the linear coefficients of *w*_1_(*<u>x</u>*) as *w*_1,1_ and *w*_2,1_. We have only linear terms contributing from the terms 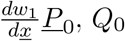 and 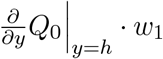, hence

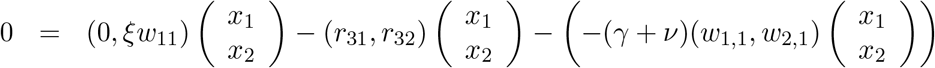

hence for the coefficients of *x*_1_

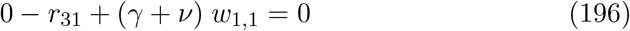

giving

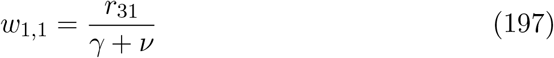

and for the coefficients of *x*_2_

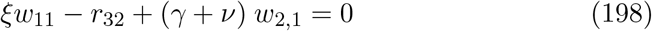

giving

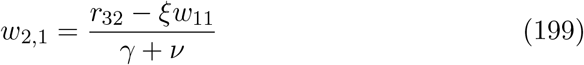

#### E.3.7 Calculaton of the quadratic coefficients of *w*_1_

Finally, we collect all quadratic terms in *<u>x</u>* in Eq. (170) to determin the quadratic coefficients of *w*_1_(*<u>x</u>*) as *w*_20,1_,*w*_11,1_ and *w*_02,1_.

First we collect the quadratic terms depending on the quadratic coefficients *w*_*ij*,1_ and keep all other terms independent on *w*_*ij*,1_ in a matrix with elements *k*_*ij*_ and obtain

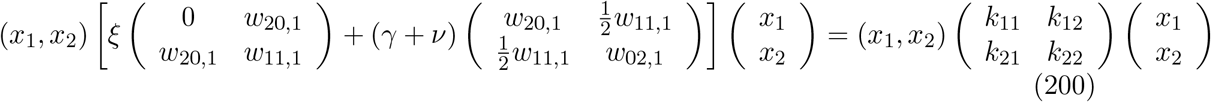

and labelling

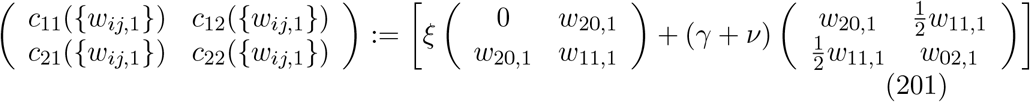

which gives the equation system for the coefficients *w*_*ij*,1_

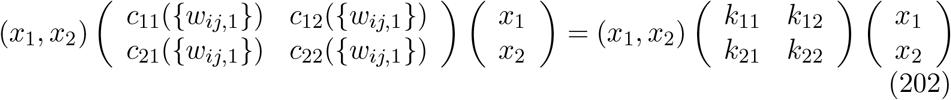

and from which we obtain the coefficients of 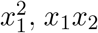 and 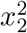 as three equations to determin *w*_20,1_,*w*_11,1_ and *w*_02,1_ as follows. For the terms with powers 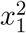 we have the coefficients’ equation given by

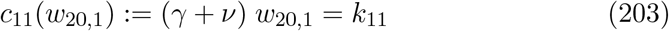

hence the first coefficient is determined by

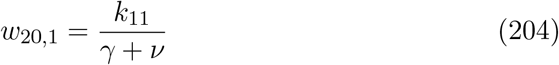

ad further for *x*_1_*x*_2_ we have the coefficients’ equation

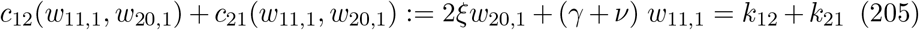

giving the second coefficient *w*_11,1_ as function of the already known *w*_20,1_ as

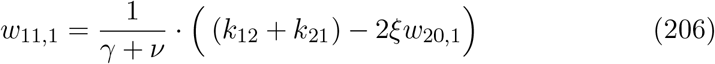

And finally for 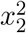 we have the coefficients’ equation

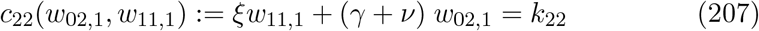

giving the final coefficient *w*_02,1_ as function of the already known *w*_11,1_ as

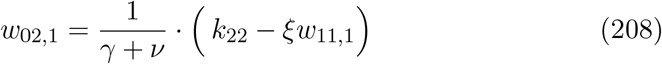

with the *k*_*ij*_ determined from Eq. (170) as

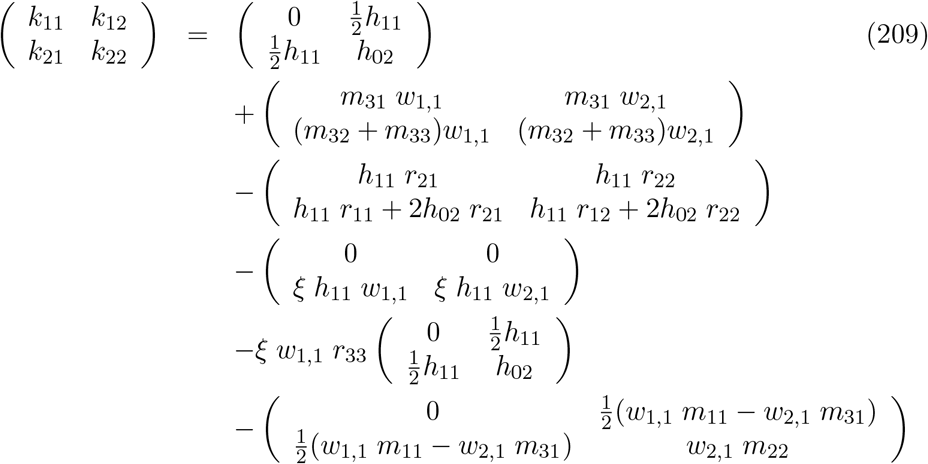

with the results for *k*_*ij*_

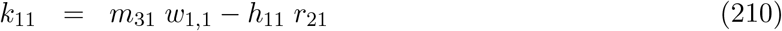

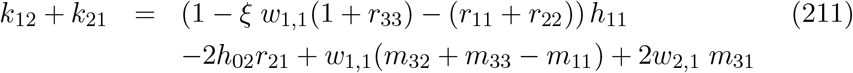

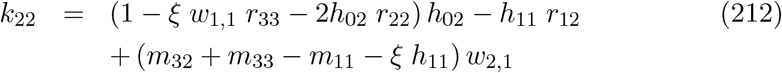

so that we have completely determined the coefficients *w*_20,1_,*w*_11,1_ and *w*_02,1_.

### E.4 Results for the coefficients of *w*_1_

We obtain for the linear part of *w*_1_(*<u>x</u>*)

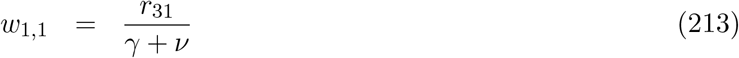

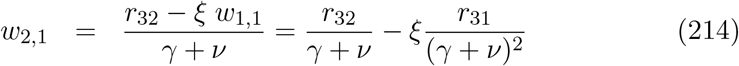

and for the quadratic part

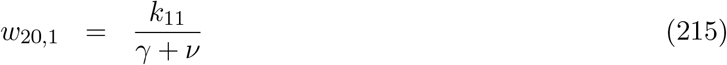

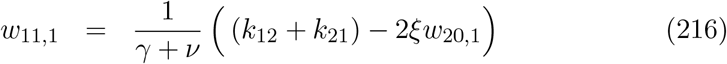

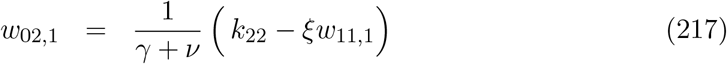

with the functions *k*_*ij*_, which depend only on model parameter and the already known coefficients of *w*(*<u>x</u>*) in linear order in *<u>x</u>*, given as

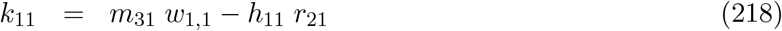

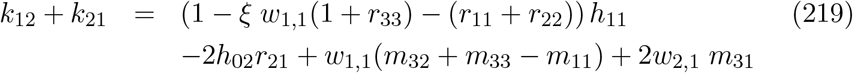

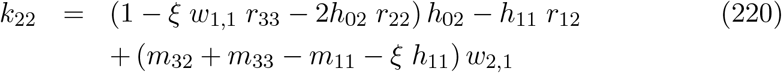

### E.5 Results for the quadratic approximation of the center manifold *z*_3_ = *z*_3_(*z*_2_, *z*_1_) in next to leading order in *µ*

In original coordinates *z*_3_ = *z*_3_(*z*_2_, *z*_1_) we now have in *O*(*µ*) to determin

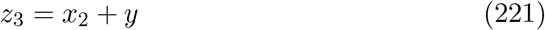

with *y* = *w*(*<u>x</u>, µ*) = *h*(*<u>x</u>*) + *µ w*(*<u>x</u>*) + *O*(*µ*^2^). Hence in quadratic order we have

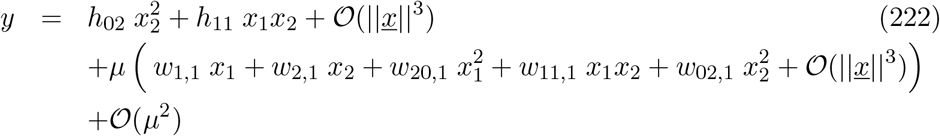

and hence for *z*_3_ we have

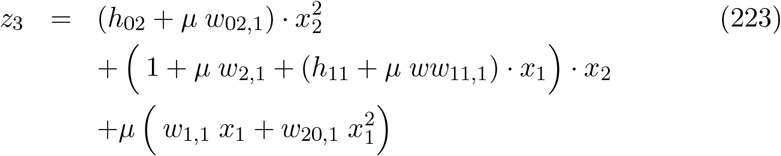

with *x*_1_ = *z*_1_ and 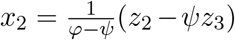 and with further calculations analogously to the zeroth order calculation of the quadratic approximation described in the previous section. From this we obtain the quadratic equation in *z*_3_ as

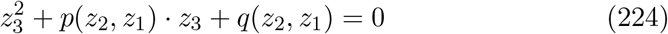

with *p*(*z*_2_, *z*_1_) and *q*(*z*_2_, *z*_1_) given by

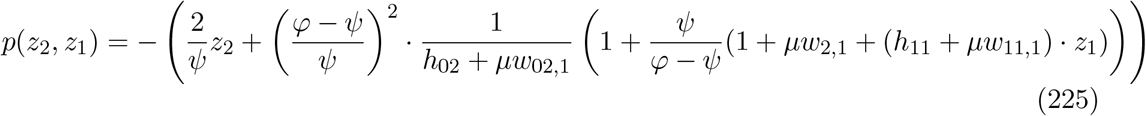

and

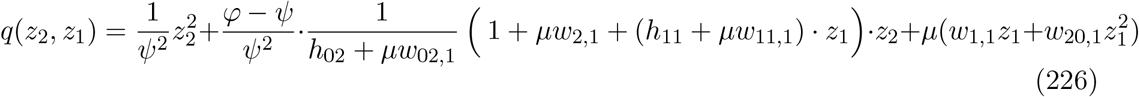

and the solution

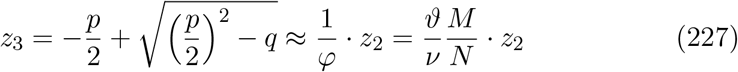

with *µ* dependent terms vanishing for *µ* going to zero, as can be seen from Eqs. (225) and (226), where *µ*-dependent terms only appear additively next to zeroth order terms.

## Footnotes

1 As an example of such calculations, here 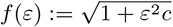 which gives in Taylor’s expansion 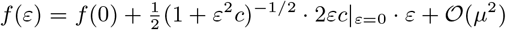, and hence *f* (*ε*) = 1 + 0 *· ε* + 𝒪 (*µ*^2^) = 1 + 𝒪 (*µ*^2^).

## Notes

### Competing Interest Statement

The authors have declared no competing interest.

### Funding Statement

M. A. has received funding from the European Unions Horizon 2020 research
and innovation programme under the Marie Sk lodowska-Curie grant agreement
No 792494. This research is also supported by the Basque Government through
BERC 2018-2021 program and by Spanish Ministry of Sciences, Innovation and
Universities: BCAM Severo Ochoa accreditation SEV-2017-0718.

### Author Declarations

IRB/oversight exemption for the research under consideration.

## References

[1] Rocha, F., Aguiar, M., Souza, M., & Stollenwerk, N. (2013) Time-scale separation and center manifold analysis describing vector-borne disease dynamics, Int. Journal. Computer Math. 90, 2105–2125.

[2] Rashkov, P., Venturino, E., Aguiar, M., Stollenwerk, N., & Kooi, B. (2019) On the role of vector modelling in a minimalistic epidemiological model, Mathematical Biosciences and Engineering 16, 4314–4338.

[3] Forgoston, E., Billings, L., & Schwartz, I.B. (2019) Model Reduction in Stochastic Environments, pp. 37–62, in Wei, W., Xiaopeng, C., & Yan, L. (eds.) Stochastic PDEs and Modelling of Multiscale Complex Systems, World Scientific, Singapore, (arXiv:1711.07842v1, 20 Nov. 2017).

[4] Forgoston, E., Billings, L., & Schwartz, I.B. (2009) Accurate noise projection for reduced stochastic epidemic models, Chaos 19, 043110.

[5] Schwartz, I.B., & Smith, H. (1983), Infinite subharmonic bifurcations in an SEIR epidemic model, J. Math. Biol. 18, 233–253.

[6] Benoît, E., Brons, M., Desroches, M., & Krupa, M. (2015) Extending the zero-derivative principle for slowfast dynamical systems, J. Zeitschrift fuer Angewandte Mathematik und Physik, 66, 2255–2270.

[7] Kaper, H.G., & Kaper, T.J. (2002) Asymptotic analysis of two reduction methods for systems of chemical reactions. Physica D: Nonlinear Phenomena, 165 66–93.

[8] Farrés, A., & Jorba, A. (2010) On the higher order approximation of the center manifold for ODEs, Discrete and Continuous Dynamical Systems, Series B 14, 977–1000.

[9] Schecter, S. (2002) Traveling-wave solutions of convection-diffusion systems by center manifold reduction, Nonlinear Analysis 49, 35–59.

[10] Kuznetsov, Y.A. (2010) Elements of applied bifurcation theory, Springer-Verlag, New York.

[11] Rocha, F., Mateus, L., Skwara, U., Aguiar, M., & Stollenwerk, N. (2016) Understanding dengue fever dynamics: a study of seasonality in vector borne disease models, International Journal of Computer Mathematics, 93, 1405–1422.

[12] Jardón-Kojakhmetov, H., Kchn, Ch., Pugliese, A. & Sensi, M. (2019) A geometric analysis of SIR, SIRS and SIRWS epidemiological models, arXiv:2002.00354v1, 4 Feb. 2020.

